# Potential pediatric tuberculosis incidence and deaths resulting from interruption in programmes supported by international health aid, 2025-2034: a mathematical modelling study

**DOI:** 10.1101/2025.05.29.25328574

**Authors:** Nicolas A Menzies, Tyler S Brown, Jeffrey W Imai-Eaton, Peter J Dodd, Ted Cohen, Leonardo Martinez

## Abstract

**Introduction:** Children experience elevated risks of developing and dying from tuberculosis (TB). We estimated the additional pediatric TB cases and deaths that could occur over 2025-2034 if programmes supported by United States bilateral health aid and the Global Fund to Fight AIDS, Tuberculosis, and Malaria (Global Fund) are discontinued.

**Methods:** We collated data on funding sources for TB and HIV programs in low- and middle-income countries and constructed scenarios representing reductions in health aid from 2025. Using calibrated transmission-dynamic models for 130 countries, we projected the discontinuation of TB and HIV treatment services under several funding reduction scenarios, and how this would affect pediatric TB exposure and treatment access. We projected pediatric TB incidence and mortality over 2025-2034 to calculate the impact of funding reductions.

**Results:** Compared to maintenance of pre-2025 service levels, withdrawal of services currently supported by US bilateral health aid was projected to result in an additional 2.5 million (95% uncertainty interval: 1.8–3.3) pediatric TB cases and 340,000 (240,000–460,000) deaths over 2025-2034. Withdrawal of US support to the Global Fund and reduction in non-US contributions was projected to result in an additional million 8.9 (6.9–11.5) pediatric TB cases and 1.5 million (1.1–2.0) deaths, more than double the number expected with continued service levels. Impacts were greatest in Sub-Saharan Africa and South-East Asia. Restoration of services in 2026 led to a substantially smaller number of additional deaths.

**Findings:** Without actions to restore discontinued services, cuts to health aid for TB and HIV programs could result in large numbers of childhood TB deaths over the next decade.

## Introduction

For most of the last 100 years, tuberculosis (TB) has been the greatest single cause of infectious disease deaths.^1^ Over recent decades, efforts to combat TB have reduced incidence rates and saved millions of lives across high-burden countries.^2^ These achievements are the product of a partnership between domestic governments, civil society, international funders, and high-income country governments that have made ongoing investments to establish and sustain TB services.

Until recently, the United States (US) was a global leader in initiatives to combat TB.^3^ US bilateral health aid for TB has been delivered predominantly through the United States Agency for International Development (USAID) and is estimated to have prevented over 75 million TB deaths in partnership with domestic governments and other funders.^4^ The United States has also been the largest single donor to the Global Fund to Fight AIDS, Tuberculosis, and Malaria (Global Fund) since its establishment in 2002, responsible for one-third of all contributions.^5^ In addition to TB-specific funding, the US President’s Emergency Plan for AIDS Relief (PEPFAR) has supported efforts to combat HIV since 2003, reversing HIV epidemics that had caused a major surge in TB cases and deaths across sub-Saharan Africa in the early 2000s.^6,7^ In early 2025, soon after the beginning of the second Trump presidential administration, the US government sharply pulled back support to these health aid programs. USAID was effectively disestablished over several months,^8,9^ and most PEPFAR activities were stalled.^10,11^ The United States also withdrew from the World Health Organization,^12,13^ and there are concerns that it will withdraw its support to the Global Fund, which begins its 8th Replenishment Cycle in late 2025.

The effect of these health aid cuts is likely to be substantial, with impacts across many countries, diseases, and population groups. Childhood TB is a condition that could be particularly sensitive to the interruption of services caused by funding cuts. Young children have the highest age-specific risks of developing TB if exposed to *M. tuberculosis* infection,^14,15^ and those who develop TB are at high risk of death without prompt treatment.^16^ Cuts to TB treatment services are expected to be followed by a rise in population-level TB incidence and prevalence. This effect could be compounded by cuts to HIV programs, as individuals with untreated HIV are highly susceptible to developing TB.^17^ For children in affected countries, this implies a potentially rapid increase in TB infection risks, at the same time that funding cuts interrupt access to diagnosis and treatment services.

In this study, we evaluated the potential impact of cuts to U.S. bilateral health aid on pediatric tuberculosis incidence and mortality in low- and middle-income countries, and the numbers of additional children that could develop and die from TB if the United States and other donors suspend or reduce their support to the Global Fund. We conducted this study for 130 countries over a 10-year period (2025-2034), to estimate the global impact of service disruptions under different funding scenarios and identify the countries and regions in which the effect of aid cuts could be most acute.

## Methods

### Population and setting

The population of interest was infants and children (0-14 years of age) living in low-and middle-income countries (defined by World Bank classifications)^18^ during 2025-2034. We excluded one country (Kosovo) due to a lack of adequate data. The remaining 130 countries represent 99.5% of global pediatric TB incidence.^19^ To create the study population we used UN Population Division estimates of annual births in each country,^20^ and assumed that without TB these individuals would face general population mortality rates.^20^

### Modelling framework

Details on the mathematical modelling approach are provided in the Supplement. In summary, the model simulated TB infection, disease, and mortality for successive birth cohorts in each country until 15 years of age, based on previous modelling approaches.^21–23^ Rates of TB exposure were generated using a previously published transmission-dynamic model of TB in the overall population calibrated to each study country.^24,25^ Risks of TB progression and mortality varied by age, malnutrition, BCG vaccination, HIV status, and receipt of TB treatment. **Figure 1** provides a schematic of the analysis, and **Table S1** reports parameter values and sources. We confirmed the fit of transmission-dynamic models against calibration data (**Figures S1**-**S2)**, and validated model estimates of pediatric TB incidence and deaths against independent data (**Figures S3**-**S4**).^19,26^

**Figure 1:**
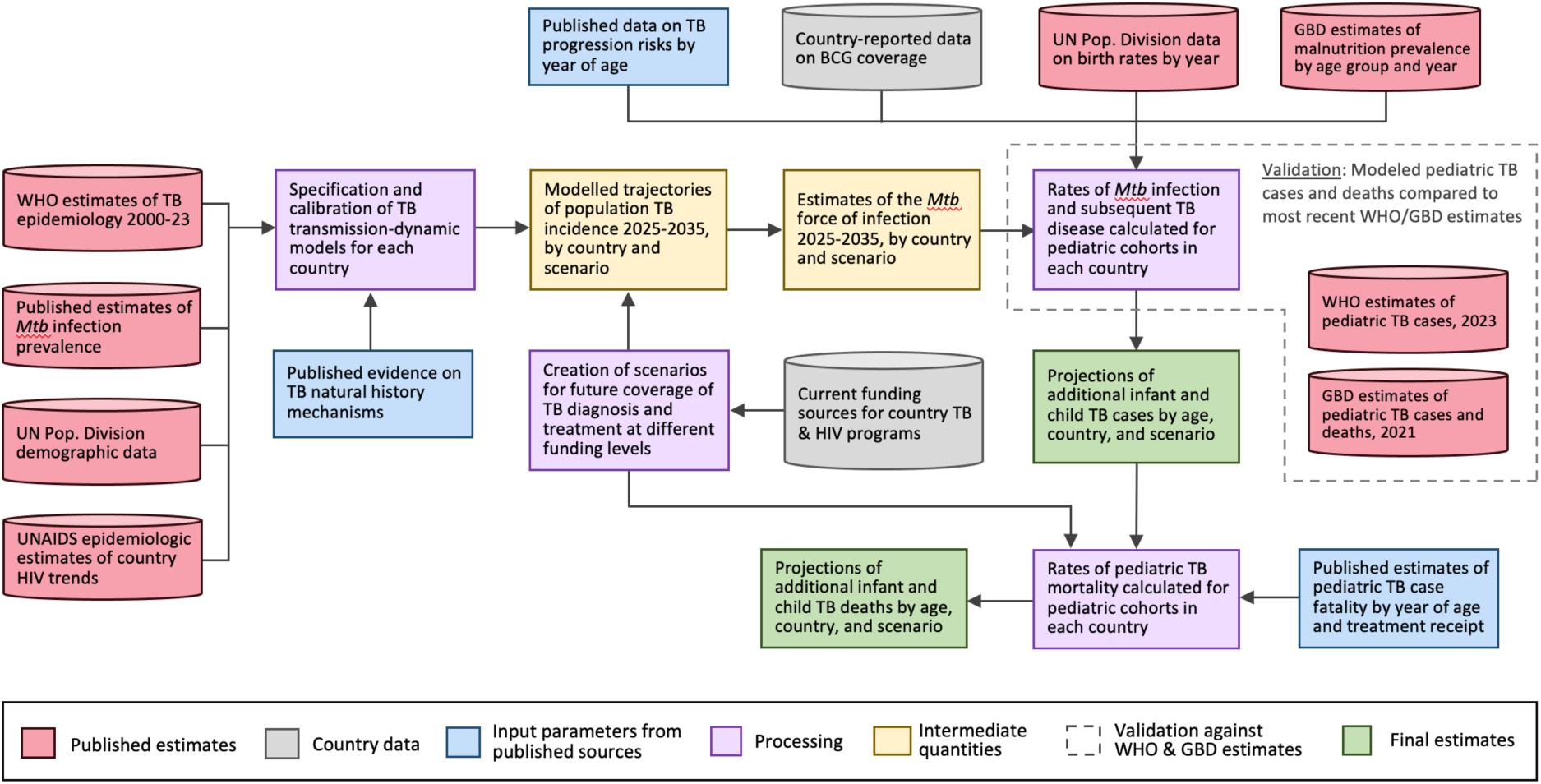
Schematic of analytic approach and data sources. TB = tuberculosis. Mtb = *Mycobacterium tuberculosis*. UN = United Nations. WHO = World Health Organization. Pop. = population. BCG = Bacillus Calmette-Guérin vaccination. GBD = Global Burden of Disease Study. Spectrum estimates represent UNAIDS Epidemiologic Estimates of country HIV trends [with the Spectrum model

### Scenarios

We extracted country-specific data on TB program expenditures by funding source (domestic, U.S. bilateral, Global Fund, other international), for the most recently available year.^19^ We extracted similar data on HIV funding.^27^ Missing or anomalous values were checked against other data sources.^28,29^ We assumed these values represented pre-2025 funding shares for TB and HIV services in each country. The U.S. share of Global Fund contributions (38%) was based on reported pledges during the 7th Global Fund Replenishment Cycle.^30^ **Figures S5**-**S6** show country-specific funding shares for TB and HIV.

We created scenarios representing alternative assumptions about the level of external support to TB and HIV programs from 2025 onwards. *Scenario 1 (base-case)* assumed the uninterrupted continuation of pre-existing funding levels. *Scenario 2 (US bilateral funding cut in 2025)* assumed that US bilateral funding for TB and HIV services would be completely cut from 2025 onwards, and other funding maintained at pre-existing levels. *Scenario 3 (US Global Fund contributions cut in 2026)* was based on Scenario 2 and additionally assumed that US Global Fund contributions would be completely cut from 2026 onwards. *Scenario 4 (50% of non-US Global Fund contributions cut in 2026)* was based on Scenario 3 and additionally assumed that 50% of non-US Global Fund contributions would be cut from 2026 onwards. Under each scenario, we assumed other sources of TB and HIV funding continued at pre-2025 levels.

Evidence is emerging on how reductions in disease-specific international funding may affect TB and HIV services, with reports describing personnel layoffs, suspension of clinical services, decreased availability of diagnostic tools, and reduced resources for patient outreach and case finding.^31,32^ We represented the impact of funding changes as reductions in access to treatment for each disease (proportion of individuals with HIV receiving ART, proportion of individuals with TB diagnosed and treated). We assumed that reductions in funding would produce less-than-proportional reductions in treatment coverage, as might result from reallocation of resources to highest priority interventions, improvements in technical efficiency, and other actions within affected countries. We operationalized this as an adjustment ratio of 0.5, varied between 0.25 and 0.75. Under this approach, a 20% reduction in total funding would produce a 10% (95% interval: 5, 15) reduction in service coverage. These reductions were applied separately for each country and disease. Using these scenario definitions, we simulated future pediatric TB cases and deaths for each country and scenario over the 2025-2034 period.

### Statistical methods

We incorporated uncertainty in parameters defining TB epidemiology and how service coverage responded to changes in funding levels. We calibrated the transmission-dynamic TB models with a Hamiltonian Monte Carlo approach,^33^ producing a sample of 1000 trajectories for the TB force of infection over the study period by country and scenario. For other model inputs we created probability distributions representing uncertainty in each parameter (**Table S1**) and sampled 1000 parameter sets using a Latin hypercube design. We simulated pediatric TB outcomes for each of these 1000 sets of model inputs. Results were calculated as the mean value of each outcome across simulations, and 95% uncertainty intervals created as the 2.5^th^ and 97.5^th^ percentiles of the simulated results. Analyses were conducted in R (v4.4.3) using the rstan (v2.32.7) and Rcpp (v1.0.14) packages.^34–36^

### Sensitivity analyses

We calculated partial rank correlation coefficients quantifying the relationship between individual parameters and total TB deaths 2025-2034 under Scenario 4. We also assessed two additional scenarios: a *low-impact scenario* in which US bilateral funding is cut for the 12 months of 2025 but then restored to full funding in 2026, and a *high-impact scenario*, which was based on Scenario 4 but with treatment coverage reduced proportional to funding reductions for each disease.

## Results

### Base-case projections

We estimated there were 1.23 million (95% uncertainty interval: 1.01–1.50) pediatric TB cases in low and middle-income countries in 2024, and 118 thousand (96–145) pediatric TB deaths. Under pre-2025 service coverage levels (Scenario 1) we projected a slow decline in pediatric TB over future years, with 1.06 thousand (0.85–1.33) pediatric TB cases and 105 thousand (83–133) pediatric TB deaths projected for 2034 respectively (average annual decline 1.5% [0.4–2.5] and 1.2% [0.2–2.3], respectively). Total projected pediatric TB cases and deaths over 2025-2034 were 11.3 million (9.2–14.0) and 1.10 million (0.89–1.38), respectively.

### Impact of interrupted TB and HIV service delivery from reduced funding

Under the reduced funding scenarios, children were projected to face progressively increasing risks of TB exposure (**Figure S7**), with those developing TB less likely to receive effective diagnosis and treatment (**Table S2**). As a consequence, pediatric TB cases and deaths were projected to rise progressively under these scenarios (**Figure 2**). For Scenario 2, representing the impact of US bilateral funding cuts from 2025 onwards, TB cases and deaths were projected to rise to 1.52 million (1.23–1.91) and 161 thousand (127–205) by 2034 (average annual increase 2.1% [0.9–3.5] and 3.1% [1.8–4.7], respectively). For Scenario 3, representing the additional impact of the United States cutting future Global Fund contributions, TB cases and deaths rose to 2.28 million (1.84–2.86) and 277 thousand (216–354) by 2034 (average annual increase 6.4% [4.9–8.1] and 8.9% [7.1–11.0] respectively). For Scenario 4, representing the additional impact of future Global Fund contributions from non-US sources falling by 50%, TB cases and deaths rose to 3.01 million (2.42–3.78) and 411 thousand (316–541) by 2034 (average annual increase 9.4% [7.7–11.4] and 13.2% [11.0–15.8] respectively). **Table 1** shows total projected pediatric TB cases and deaths over the 10-year study period for each scenario. As compared to the base-case scenario, total pediatric TB deaths for 2025-2034 were projected to rise by 31% (24–40), 84% (67–106), and 134% (110–178), under Scenarios 2, 3, and 4 respectively (**Table S3**).

**Figure 2:**
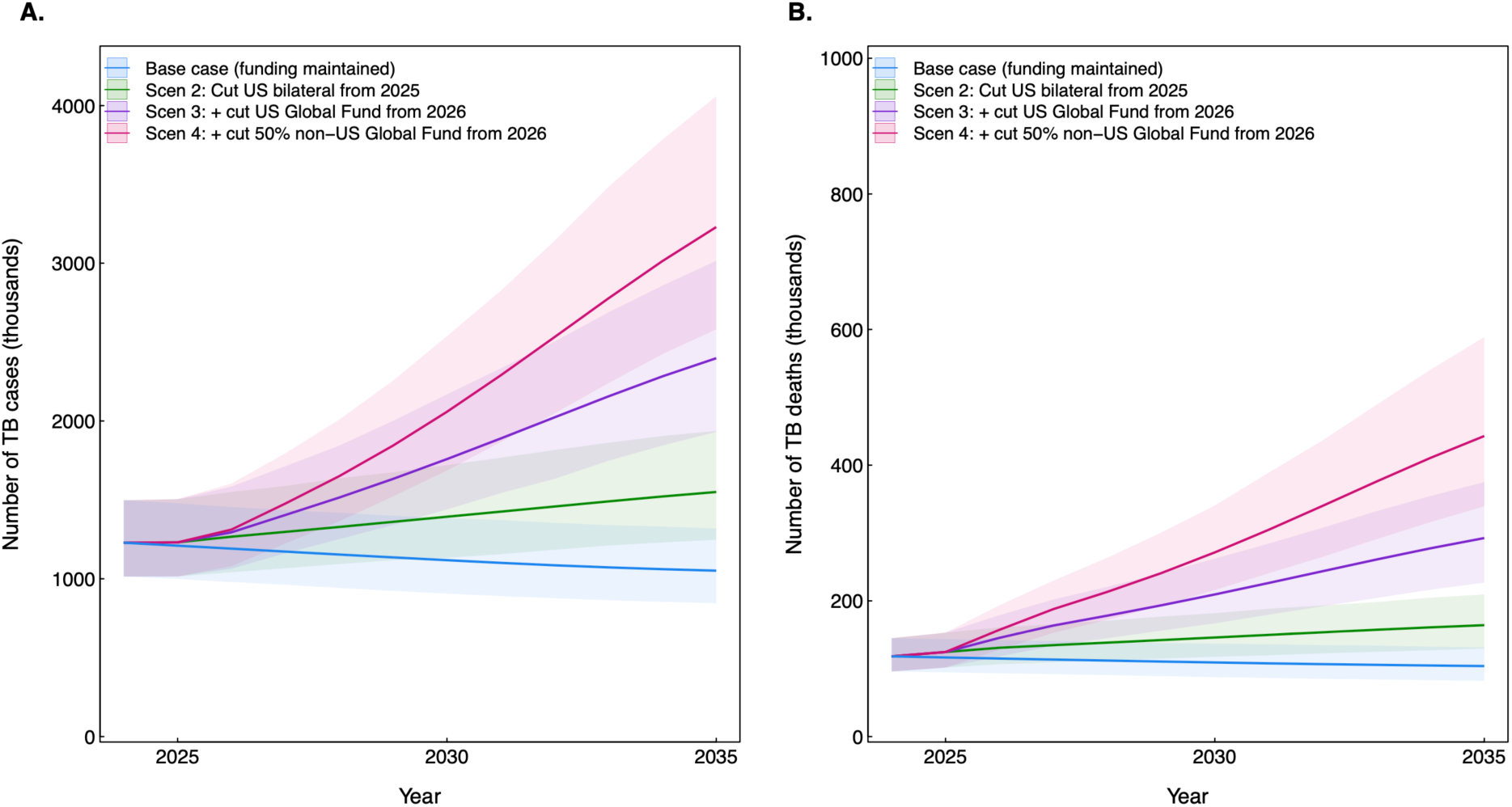
Annual pediatric TB cases (Panel A) and deaths (Panel B) in 130 low- and middle-income countries projected under each funding scenario, 2025-2034. Scen = scenario. TB = tuberculosis. US = United States. Scenarios 2-4 represent progressively greater funding cuts, such that Scenario 4 includes 100% reductions in US bilateral funding in 2025 and Global Fund support in 2026, and 50% reductions in non-US Global Fund support in 2026.

**Table 1:**
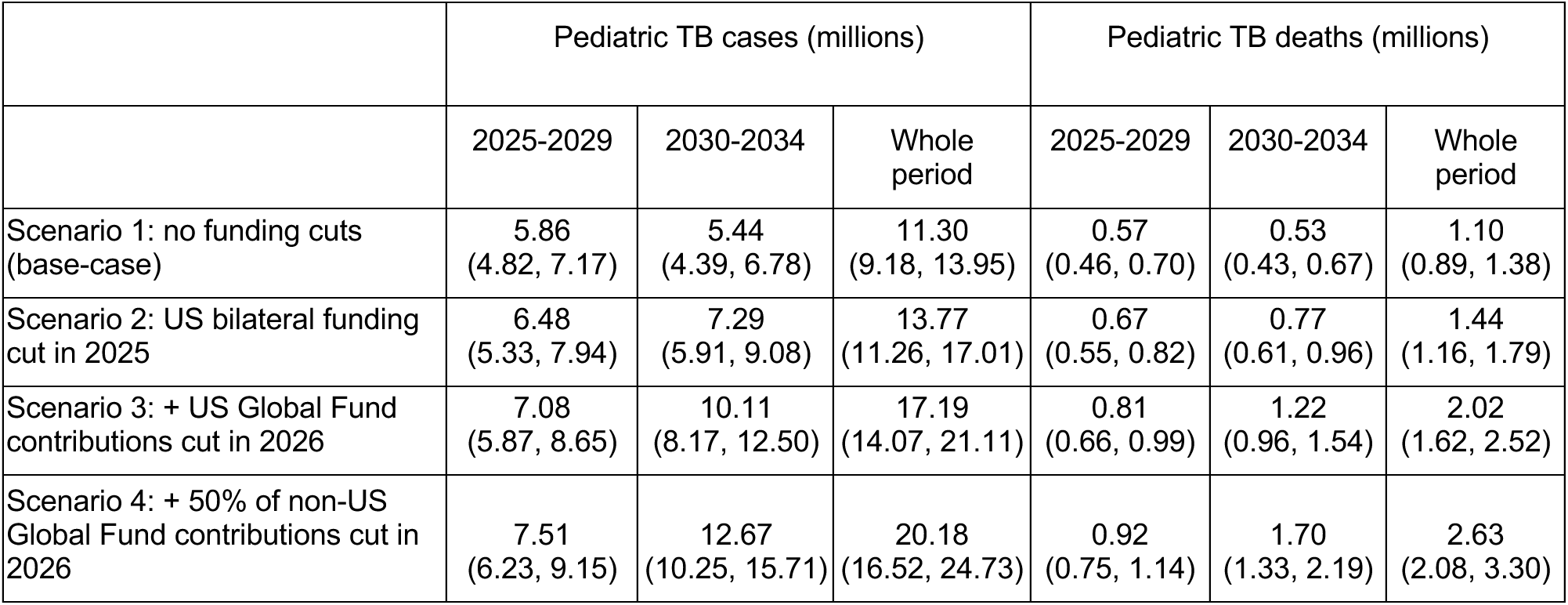
Pediatric TB cases and deaths in 130 low- and middle-income countries projected under each funding scenario, 2025-2034. TB = tuberculosis. US = United States. Scenarios 2-4 represent progressively greater funding cuts, such that Scenario 4 includes 100% reductions in US bilateral funding in 2025 and Global Fund support in 2026, and 50% reductions in non-US Global Fund support in 2026.

|  | Pediatric TB cases (millions) |  |  | Pediatric TB deaths (millions) |  |  |
| --- | --- | --- | --- | --- | --- | --- |
|  | 2025-2029 | 2030-2034 | Whole period | 2025-2029 | 2030-2034 | Whole period |
| Scenario 1: no funding cuts (base-case) | 5.86<br>(4.82, 7.17) | 5.44<br>(4.39, 6.78) | 11.30<br>(9.18, 13.95) | 0.57<br>(0.46, 0.70) | 0.53<br>(0.43, 0.67) | 1.10<br>(0.89, 1.38) |
| Scenario 2: US bilateral funding cut in 2025 | 6.48<br>(5.33, 7.94) | 7.29<br>(5.91, 9.08) | 13.77<br>(11.26, 17.01) | 0.67<br>(0.55, 0.82) | 0.77<br>(0.61, 0.96) | 1.44<br>(1.16, 1.79) |
| Scenario 3: + US Global Fund contributions cut in 2026 | 7.08<br>(5.87, 8.65) | 10.11<br>(8.17, 12.50) | 17.19<br>(14.07, 21.11) | 0.81<br>(0.66, 0.99) | 1.22<br>(0.96, 1.54) | 2.02<br>(1.62, 2.52) |
| Scenario 4: + 50% of non-US Global Fund contributions cut in 2026 | 7.51<br>(6.23, 9.15) | 12.67<br>(10.25, 15.71) | 20.18<br>(16.52, 24.73) | 0.92<br>(0.75, 1.14) | 1.70<br>(1.33, 2.19) | 2.63<br>(2.08, 3.30) |

### Distribution of health impacts

Additional TB cases and deaths projected under the reduced funding scenarios were concentrated in countries and regions with high current TB incidence, low current TB treatment coverage, and a high share of current TB and HIV program funding from non-domestic sources (**Table 2**). Consequently, impacts were inversely related to country income level, with relative increases in pediatric cases and deaths highest for low-income countries, followed by lower-middle and upper-middle income countries respectively (**Table S4**)

**Table 2:** Additional pediatric TB cases and deaths under alternative funding scenarios as compared to continued funding, 2025-2034, by country grouping. TB = tuberculosis. US = United States. Scenario 2: 100% of US bilateral funding cut in 2025. Scenario 3: 100% of US bilateral funding cut in 2025, plus 100% of US Global Fund contributions cut in 2026. Scenario 4: 100% of US bilateral funding cut in 2025, plus 100% of US and 50% of non-US Global Fund contributions cut in 2026. Income groups defined according to World Bank categorizations. High TB burden group includes the 30 countries identified as having high TB burden by WHO. High non-domestic funding group represents countries with >80% non-domestic funding for TB or HIV programs.

|  | Thousands of pediatric TB cases |  |  | Thousands of pediatric TB deaths |  |  |
| --- | --- | --- | --- | --- | --- | --- |
|  | Scenario 2 | Scenario 3 | Scenario 4 | Scenario 2 | Scenario 3 | Scenario 4 |
| Total | 2,473<br>(1,806, 3,336) | 5,888<br>(4,580, 7,599) | 8,885<br>(6,884, 11,454) | 337<br>(244, 459) | 921<br>(696, 1,225) | 1,524<br>(1,138, 2,031) |
| WHO region |  |  |  |  |  |  |
| *Eastern Mediterranean Region* | 65<br>(22, 147) | 733<br>(352, 1,387) | 1,340<br>(633, 2,594) | 8.2<br>(2.6, 20.8) | 117<br>(54, 241) | 239<br>(106, 505) |
| *African Region* | 1,425<br>(944, 2,111) | 2,926<br>(2,046, 4,190) | 4,244<br>(3,018, 6,026) | 205<br>(132, 307) | 484<br>(324, 722) | 770<br>(522, 1,152) |
| *European Region* | 2.0<br>(0.8, 4.0) | 8.2<br>(4.7, 13.7) | 14<br>(8, 24) | 0.30<br>(0.12, 0.60) | 1.1<br>(0.6, 1.9) | 1.9<br>(1.1, 3.2) |
| *Region of the Americas* | 1.6<br>(0.7, 3.3) | 26<br>(15, 47) | 49<br>(27, 90) | 0.12<br>(0.05, 0.26) | 3.4<br>(1.8, 6.3) | 7.2<br>(3.5, 14.2) |
| *South-East Asian Region* | 822<br>(461, 1,420) | 1,809<br>(1,071, 2,942) | 2,655<br>(1,540, 4,280) | 98<br>(55, 164) | 253<br>(140, 409) | 406<br>(218, 678) |
| *Western Pacific Region* | 157<br>(65, 319) | 386<br>(203, 686) | 584<br>(322, 995) | 25<br>(10, 52) | 63<br>(31, 117) | 100<br>(51, 180) |
| Income group |  |  |  |  |  |  |
| *Low income* | 1,119<br>(668, 1,769) | 2,380<br>(1,569, 3,602) | 3,484<br>(2,309, 5,179) | 147<br>(84, 243) | 370<br>(229, 603) | 604<br>(386, 986) |
| *Lower middle income* | 1,222<br>(819, 1,848) | 2,884<br>(2,066, 3,997) | 4,355<br>(3,114, 6,105) | 170<br>(114, 244) | 448<br>(310, 632) | 733<br>(506, 1,047) |
| *Upper middle income* | 132<br>(65, 250) | 624<br>(275, 1,274) | 1,046<br>(449, 2,156) | 19<br>(9, 38) | 104<br>(43, 223) | 188<br>(73, 418) |
| TB burden category |  |  |  |  |  |  |
| *High TB burden* | 2,345<br>(1,681, 3,193) | 5,167<br>(3,889, 6,876) | 7,624<br>(5,720, 10,208) | 320<br>(228, 441) | 801<br>(576, 1,110) | 1,290<br>(916, 1,795) |
| *Not high TB burden* | 128<br>(76, 216) | 721<br>(542, 979) | 1,261<br>(961, 1,674) | 17<br>(10, 29) | 120<br>(89, 165) | 234<br>(173, 321) |
| Funding category |  |  |  |  |  |  |
| *High non-domestic funding* | 1,545<br>(1,053, 2,223) | 3,322<br>(2,375, 4,630) | 4,878<br>(3,551, 6,710) | 220<br>(143, 329) | 546<br>(374, 798) | 883<br>(610, 1,284) |
| *Non high non-domestic funding* | 928<br>(550, 1,494) | 2,565<br>(1,730, 3,748) | 4,007<br>(2,666, 5,803) | 117<br>(68, 191) | 375<br>(251, 555) | 641<br>(415, 980) |

In 42 countries with at least 80% of TB and/or HIV funding from non-domestic sources, TB cases and deaths for 2025-2034 were 120% (92–159) and 190% (137–270) higher under Scenario 4 compared to the base-case. Additional TB cases and deaths were concentrated in African and South-East Asian regions (**Figure 3**). **Table S5** reports results for individual countries.

**Figure 3:**
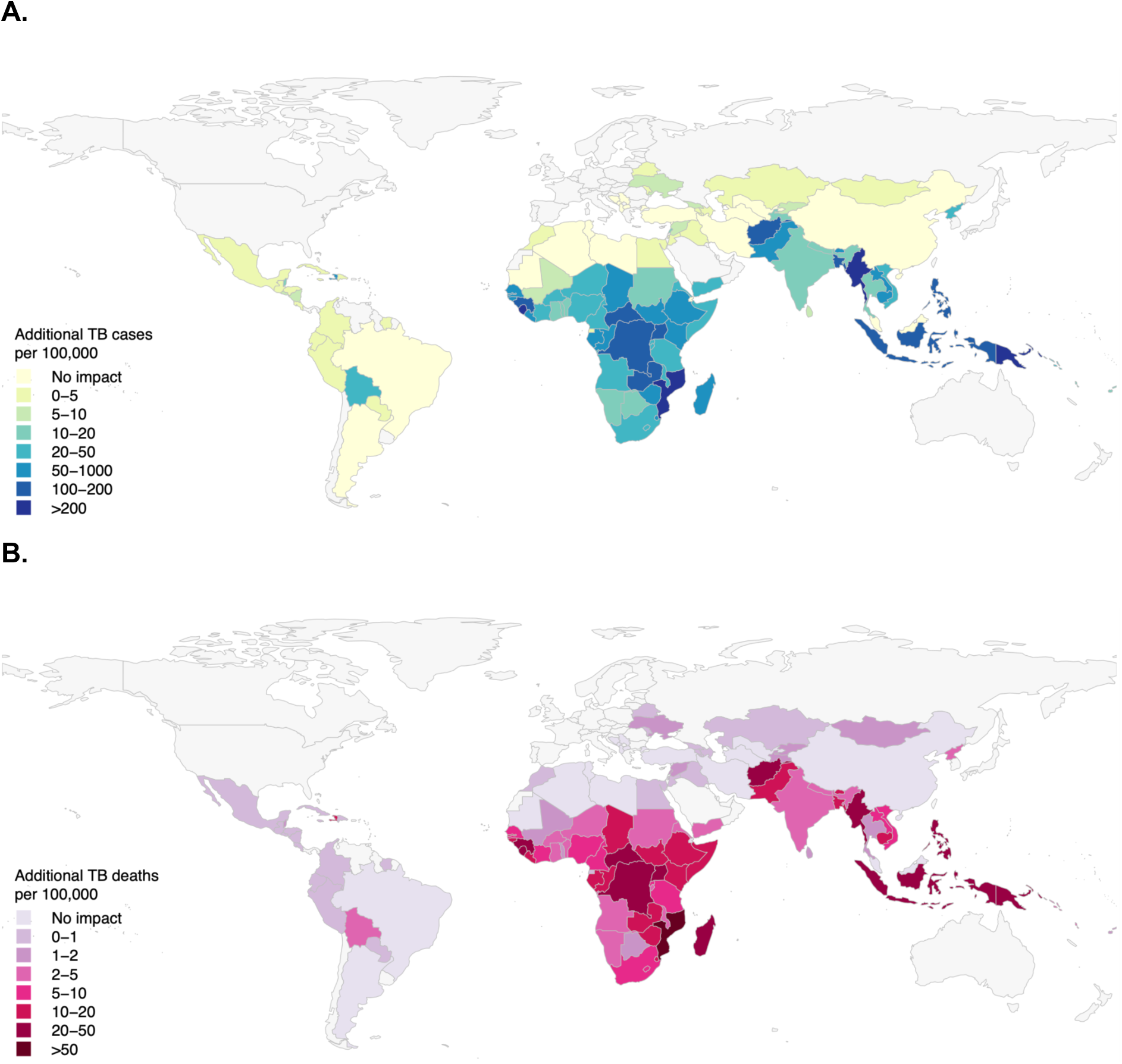
Additional TB cases (Panel A) and deaths (Panel B) under the most extreme funding reduction scenario, as compared to continued funding, 2025-2034. TB = tuberculosis. US = United States. Scenario represents 100% of US bilateral funding cut in 2025, plus 100% of US and 50% of non-US Global Fund contributions cut in 2026 (Scenario 4).

### Sensitivity analyses

The most influential inputs included parameters describing the force of infection faced by children, the sensitivity of TB and HIV service coverage to funding reductions, and the probability of TB following infectious exposure (partial rank correlation coefficients shown in **Figure S8**). **Table S6** reports results for the *low-impact and high-impact* scenarios, representing optimistic and pessimistic scenarios for the impact of funding reductions. Under the *low-impact* scenario an additional 39 thousand (29–51) pediatric TB deaths were projected, 3.5% (2.8–4.3) greater than the base-case. Under the *high-impact* scenario an additional 3.62 million (2.93–4.52) pediatric TB deaths were projected, 330% (279–385) greater than the base-case.

## Discussion

In this global analysis of pediatric tuberculosis incidence and mortality, disruption of TB and HIV services arising from broad-scale cuts to US health aid were projected to result in substantial numbers of children developing and dying from TB over 2025-2034. Under a conservative scenario in which only US bilateral health aid is discontinued, approximately 2.5 million additional pediatric tuberculosis cases and 340,000 pediatric tuberculosis deaths are expected. If the United States and other countries reduce support for the Global Fund a further 3–7 million additional cases and 0.6–1.2 million additional child deaths could occur, reflecting the central role of this multilateral funder in supporting TB and HIV services in the most affected countries. These findings suggest catastrophic impacts that could result from health aid cuts, which will grow over time as the duration of service disruptions increases.

Our analysis suggests that the impacts of recent funding cuts will be most apparent in the WHO African and South-East Asian regions. In these regions many countries had high TB and HIV burden before 2025, face substantial health system constraints, and had a heavy reliance on health aid to support current services. We projected the Americas and European regions would be largely unaffected. Some of the countries projected to incur the greatest impacts included those with high HIV prevalence. In these countries, reductions in HIV funding contributed to the overall impact, with reduced ART access projected to produce a growing pool of persons with untreated HIV, highly susceptible to TB.

These results are consistent with other studies that investigated the potential implications of TB funding cuts.^37–40^ Compared to those analyses, this study included more countries, considered the additional implications of HIV funding cuts for TB, and focussed on an age group at high risk of TB and mortality. Nevertheless, several key findings are shared with these analyses - that the largest impacts are estimated for settings with limited capacity to respond to these challenges, and that the magnitude of harm will grow over time as the duration of the funding gap increases.

There are several potential policy implications from this analysis. Importantly, this analysis illustrates what is at stake under different levels of health aid to support TB and HIV services. Notably, the projected number of pediatric TB deaths was substantially smaller when we assumed funding would be restored after one year, with the additional deaths projected for this scenario reduced by 90% or more compared to our other scenarios. This demonstrates that the prompt restoration of funding, or the creation of alternative financing mechanisms to sustain current services, could greatly reduce the consequences of funding cuts. In parallel with actions to restore or replace lost funding, efforts to identify efficiencies, prioritize the highest impact interventions, and target services to the highest need populations will play a key role in limiting service disruptions and their health consequences. In our analyses we assumed that improvements in technical and allocative efficiency could produce a less-than-proportional reduction in service coverage following funding cuts. How to achieve these efficiencies is an urgent research priority.

Strengths of our analysis include the detailed country-specific estimates of risk, the consideration of both TB and HIV funding cuts, and inclusion of all LMIC to estimate how funding cuts could impact global outcomes. Moreover, prior analyses have not considered cuts to Global Fund contributions, yet these contribution levels were not set as of mid-2025, and in our analyses reductions in Global Fund contributions had the largest consequences for future outcomes. There are also several limitations. First, we examined a small number of funding scenarios. While these reflect a range of funding levels, the timing and magnitude of actual funding cuts will differ from those examined. Second, we assumed the same relationship between funding reductions and service coverage for all countries. In reality, these relationships may vary between countries and over time, as policy and clinical practice respond to new resource constraints. For both these reasons, impact estimates will need to be updated as new evidence becomes available.^41^ Third, there is substantial uncertainty about many of the other determinants of pediatric TB outcomes, reflected in wide uncertainty intervals for country-specific results. Finally, we didn’t consider all potential mechanisms through which funding cuts could affect pediatric TB deaths. Increases in malnutrition,^42^ reduced BCG coverage,^43^ and broad health system failure are each possible, and could further increase pediatric TB deaths.

Taken together, these results suggest that unless reversed or mitigated through alternative mechanisms, cuts to global health aid will have amongst the largest impacts on the pediatric tuberculosis epidemic of any single event of the past three decades, reversing global declines in pediatric TB that have been achieved during this time.

## Data Availability

All data produced in the present study are available upon reasonable request to the authors.

