## Supplement for "Potential pediatric tuberculosis incidence and deaths resulting from interruption in programmes supported by international health aid, 2025-2034: a mathematical modelling study"

### Supplementary methods

Pediatric TB model

We simulated successive monthly birth cohorts in each country, from birth to 15 years of age. Cohorts were simulated from 2010 (to produce a full cross-section of the pediatric population in 2025) until the end of 2034. Rates of TB exposure were allowed to vary by age, country, and calendar month (see '*TB force of infection*' section). Assumptions around risks of TB incidence and mortality were based on approaches adopted in previous modeling studies,[1-3] with the risks of TB for exposed children varying by age, prevalence of malnutrition, and BCG vaccination coverage. Case fatality rates for children developing TB were assumed to vary by age and receipt of TB treatment. For the subset of countries with general population HIV prevalence >1% or HIV prevalence among TB cases >5% ('co-prevalent HIV' countries, n=63), we additionally stratified the pediatric population by HIV status (HIV uninfected, HIV infected receiving ART, HIV infected not receiving ART), with the fraction of the pediatric population in each HIV stratum based on Spectrum model projections for each country[4] submitted by countries to UNAIDS as part of UNAIDS 2024 Global Epidemiologic Estimates. In these countries the risks of TB incidence and mortality were stratified by HIV status in addition to other factors. **Table S1** reports model parameter values and sources.

TB force of infection

Rates of TB exposure for children in the study cohort were based on estimates of the TB force of infection produced by a published transmission-dynamic model of TB in the overall population for each study country.[5,6] This model was recalibrated to the most recent TB epidemiological estimates for each setting,[7,8] with natural history assumptions matching published ranges for TB modelling analyses.[9] For the 63 countries with co-prevalent HIV, the modelled population was distributed across a set of six HIV strata representing HIV status, immune function (categorized by CD4 cell count) and ART receipt. The proportion of the population in each HIV stratum (by age group and year) was based on Spectrum projections.[4] **Figures S1**-**S2** compare epidemiological estimates from the transmission-dynamic model against available data.

Model validation

The modeling framework was used to simulate pediatric TB incidence and deaths in each country over recent years. These results were compared to the most recent estimates produced by the WHO (for TB incidence) and the Global Burden of Disease Study (for TB incidence and TB deaths) among 0-5-year-olds and 0-14-year-olds (**Figures S3**-**S4**).[7,10]

| Input | Values (95% interval) | Distribution | Source |
|---|---|---|---|
| *Cohort demographics and risk factor prevalence* | | | |
| Number of new births | Stratified by year and country | Fixed values | UN World Population Prospects[11] |
| Background mortality rates | Stratified by age, year and country | Fixed values | UN World Population Prospects[11] |
| HIV prevalence and ART receipt | Stratified by age, year, country, and scenario | Fixed values | UNAIDS 2024 Epidemiologic Estimates, Spectrum HIV files[4] |
| BCG vaccination coverage | Stratified by year and country | Fixed values | WHO/UNICEF[12] |
| Prevalence of malnutrition | Stratified by age, year and country | Fixed values | Institute for Health Metrics and Evaluation[10] |
| *Risks of TB exposure* | | | |
| TB force of infection | Stratified by year, country, and scenario | Sampled values from calibration | Transmission-dynamic models* |
| Effective contact rate, relative to adult population, age [0, 5) years | 0.20 (0.10, 0.40) | Gamma(6.66,33.3) | Zelner et al 2014[13] |
| Effective contact rate, relative to adult population, age [5, 10) years | 0.30 (0.20, 0.50) | Gamma(15.2, 50.6) | Zelner et al 2014[13] |
| Effective contact rate, relative to adult population, age [10, 15) years | 0.40 (0.30, 0.60) | Gamma(27.1, 67.9) | Zelner et al 2014[13] |
| *Risks of TB following TB exposure* | | | |
| Probability of TB following infectious exposure, age [0, 1) years | 0.50 (0.25, 0.75) | Beta(6.92, 6.92) | Dodd et al, 2014[2] |
| Probability of TB following infectious exposure, age [1, 2) years | 0.25 (0.125, 0.50) | Beta(4.69, 14.1) | Dodd et al, 2014[2] |
| Probability of TB following infectious exposure, age [2, 5) years | 0.05 (0.025, 0.10) | Beta(6.27, 119) | Dodd et al, 2014[2] |
| Probability of TB following infectious exposure, age [5, 10) years | 0.02 (0.01, 0.04) | Beta(6.50, 319) | Dodd et al, 2014[2] |
| Probability of TB following infectious exposure, age [10, 15) years | 0.15 (0.075, 0.30) | Beta(5.49, 31.1) | Dodd et al, 2014[2] |
| Odds ratio of TB with BCG vaccination, latitude <20° | 0.77 (0.49, 1.00) | Gamma(34.9, 45.3) | Mangtani et al, 2014[14] |
| Odds ratio of TB with BCG vaccination, latitude 20-40° | 0.68 (0.43, 0.95) | Gamma(26.1, 38.4) | Mangtani et al, 2014[14] |
| Odds ratio of TB with BCG vaccination, latitude >40° | 0.31 (0.20, 0.45) | Gamma(23.5, 75.7) | Mangtani et al, 2014[14] |
| Odds ratio of TB with malnutrition | 4.0 (2.0, 6.0) | Gamma(15.2, 3.80) | Lonnroth 2010[15] |
| Odds ratio of TB with HIV not on ART | 20 (10, 50) | Gamma(3.67, 0.184) | Dodd et al, 2017[1] |
| Odds ratio of TB with HIV, ART vs. not on ART | 0.3 (0.21, 0.39) | Gamma(42.5, 142) | Dodd et al, 2017[1] |
| *Risks of TB mortality with TB* | | | |
| Case fatality of TB with treatment, age [0, 5) years | 0.019 (0.005, 0.071) | Beta(1.08, 56.0) | Dodd et al, 2017[1] |

| Input | Values (95% interval) | Distribution | Source |
|---|---|---|---|
| Case fatality of TB without treatment, age [0, 5) years | 0.44 (0.37, 0.51) | Beta(85.9, 111) | Dodd et al, 2017[1] |
| Case fatality of TB without treatment, age [0, 5) years, HIV infected not on ART | 0.90 (0.78, 0.97) | Beta(31.7, 3.52) | Dodd et al, 2017[1] |
| Case fatality of TB without treatment, age [0, 5) years, HIV infected on ART | 0.64 (0.30, 0.90) | Beta(5.21, 2.99) | Dodd et al, 2017[1] |
| Case fatality of TB with treatment, age [5, 15) years | 0.008 (0.003, 0.021) | Beta(2.84, 352) | Dodd et al, 2017[1] |
| Case fatality of TB without treatment, age [1, 15) years | 0.15 (0.12, 0.19) | Beta(49.9, 285) | Dodd et al, 2017[1] |
| Case fatality of TB without treatment, age [5, 15) years, HIV infected not on ART | 0.87 (0.67, 0.97) | Beta(14.6, 2.11) | Dodd et al, 2017[1] |
| Case fatality of TB without treatment, age [5, 15) years, HIV infected on ART | 0.46 (0.12, 0.83) | Beta(2.76, 3.28) | Dodd et al, 2017[1] |
| Odds ratio of TB case fatality for TB, untreated vs. treated, age [5, 15) years, HIV infected not on ART | 13.9 (5.4, 36.2) | Gamma(2.96, 0.213) | Dodd et al, 2017[1] |
| Odds ratio of TB case fatality for TB, untreated vs. treated, age [5, 15) years, HIV infected on ART | 7.8 (2.3, 27.0) | Gamma(1.38, 0.176) | Dodd et al, 2017[1] |
| *Natural history delays* | | | |
| Rate of progressing to clinical TB, for newly infected individuals, age [0, 1) years | 6.0 (3.0, 9.0) | Gamma(15.2, 2.53) | Cruz et al 2007[16] ** |
| Rate of progressing to clinical TB, for newly infected individuals, age [1, 15) years | 3.0 (1.5, 6.0) | Gamma(6.66, 2.22) | Cruz et al 2007[16] *** |
| Rate of TB mortality, for individuals developing TB, age [0, 1) years | 6.0 (3.0, 9.0) | Gamma(15.2, 2.53) | Assumed** |
| Rate of TB mortality, for individuals developing TB, age [1, 15) years | 3.0 (1.5, 6.0) | Gamma(6.66, 2.22) | Assumed*** |
| *Implications of funding reductions* | | | |
| Percentage reduction in TB and HIV service coverage per percentage reduction in funding levels | 0.50 (0.25, 0.75) | Gamma(15.2, 30.4) | Assumed |

**Table S1: Values, distributions, and sources of input data for pediatric TB model.**

* Transmission-dynamic models for each country were adapted from earlier modelling studies [cites], recalibrated to the most recent epidemiological data for each country. ** Equivalent to an average 2.0 (1.3, 4.0) month delay. *** Equivalent to an average 4.0 (2.0, 8.0) month delay. Gamma distributions parameterized as shape and rate

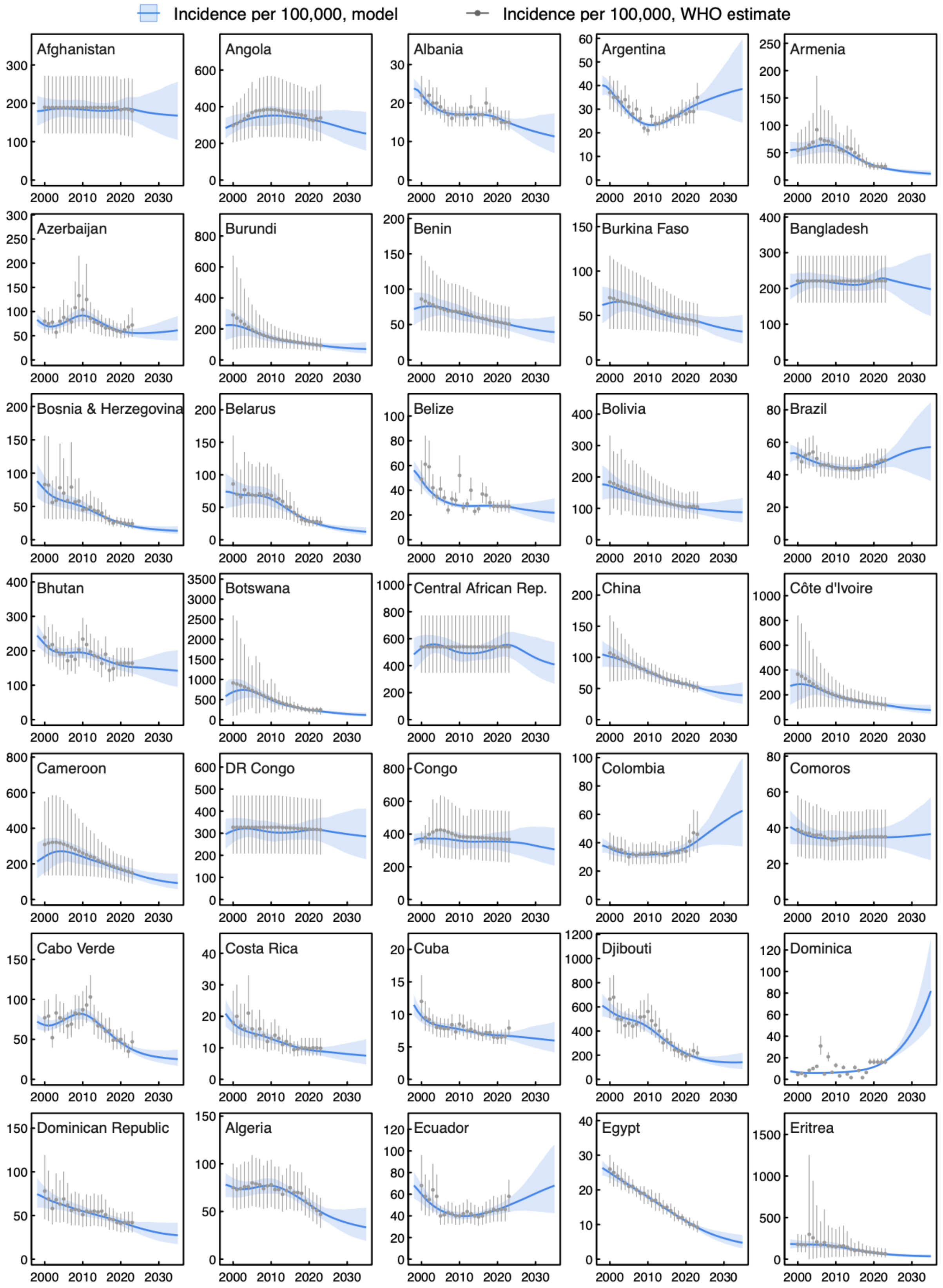

**Figure S2. Fit of country-specific transmission-dynamic models to WHO TB incidence estimates for 2000-2023, with model base-case projections to 2035.**

For WHO estimates, bars represent published uncertainty intervals. For modelled values, shaded region represents 95% uncertainty interval.

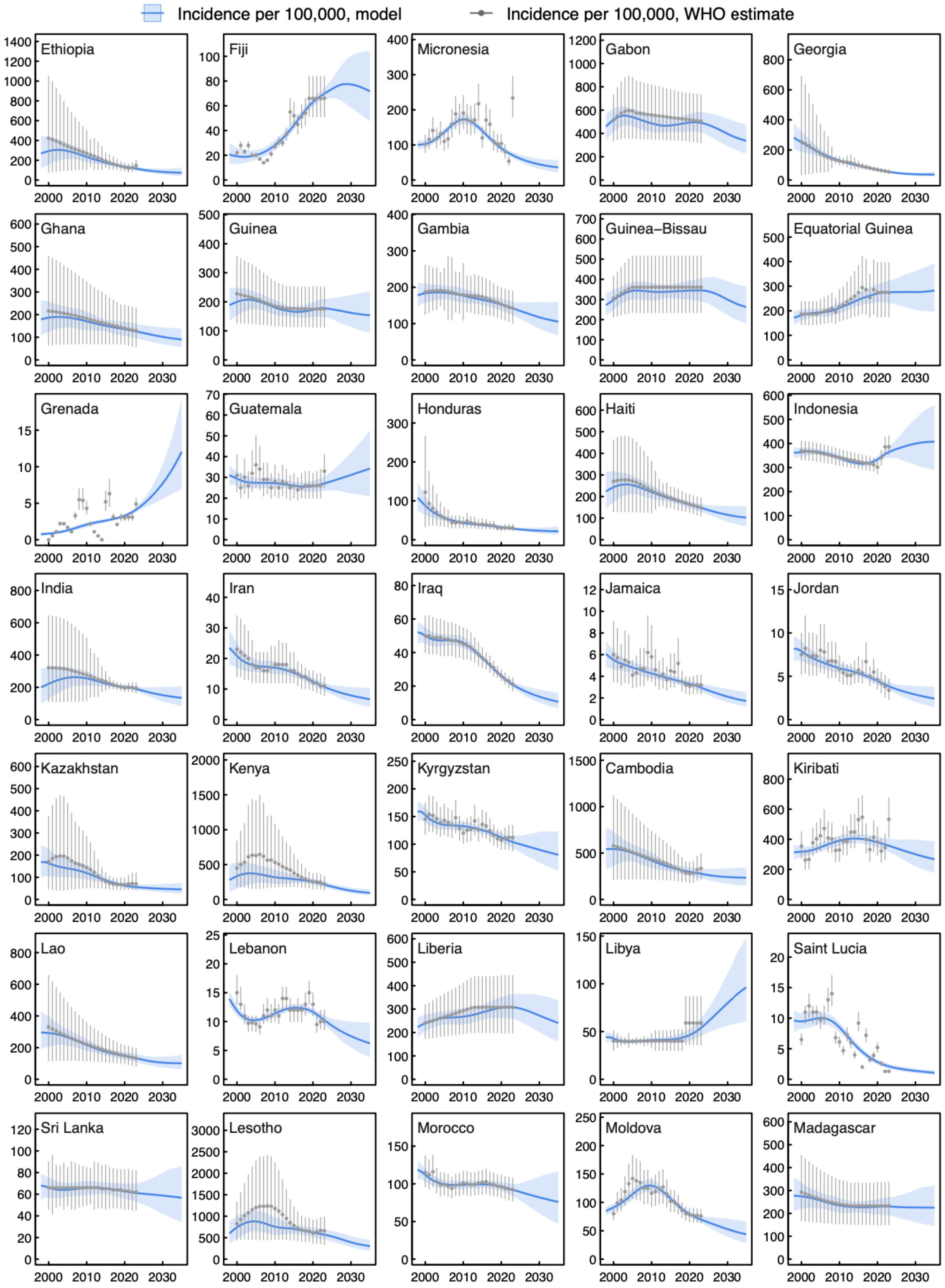

**Figure S2 continued. Fit of country-specific transmission-dynamic models to WHO TB incidence estimates for 2000-2023, with model base-case projections to 2035.**

For WHO estimates, bars represent published uncertainty intervals. For modelled values, shaded region represents 95% uncertainty interval.

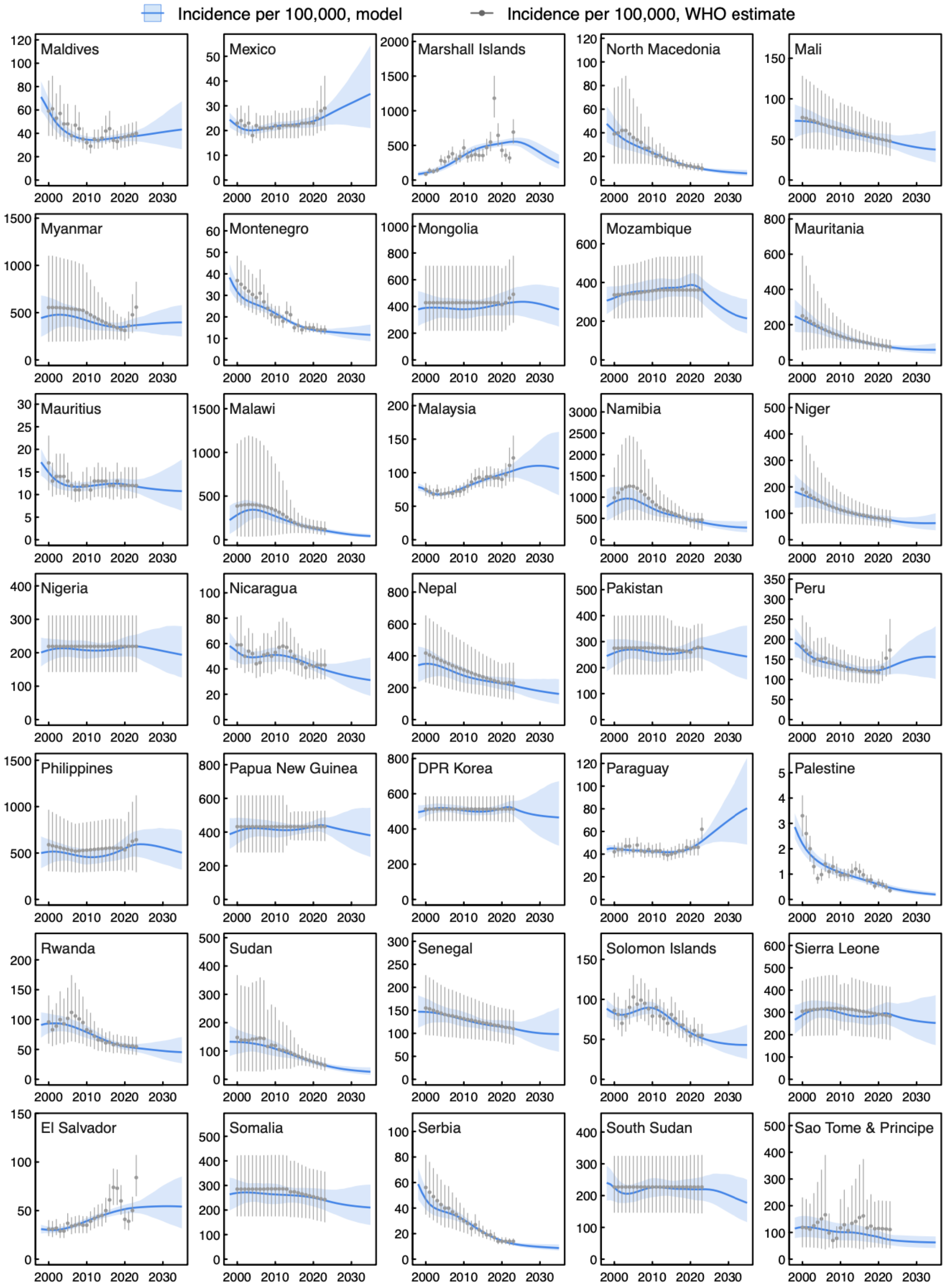

**Figure S2 continued. Fit of country-specific transmission-dynamic models to WHO TB incidence estimates for 2000-2023, with model base-case projections to 2035.**

For WHO estimates, bars represent published uncertainty intervals. For modeled values, shaded region represents 95% uncertainty interval.

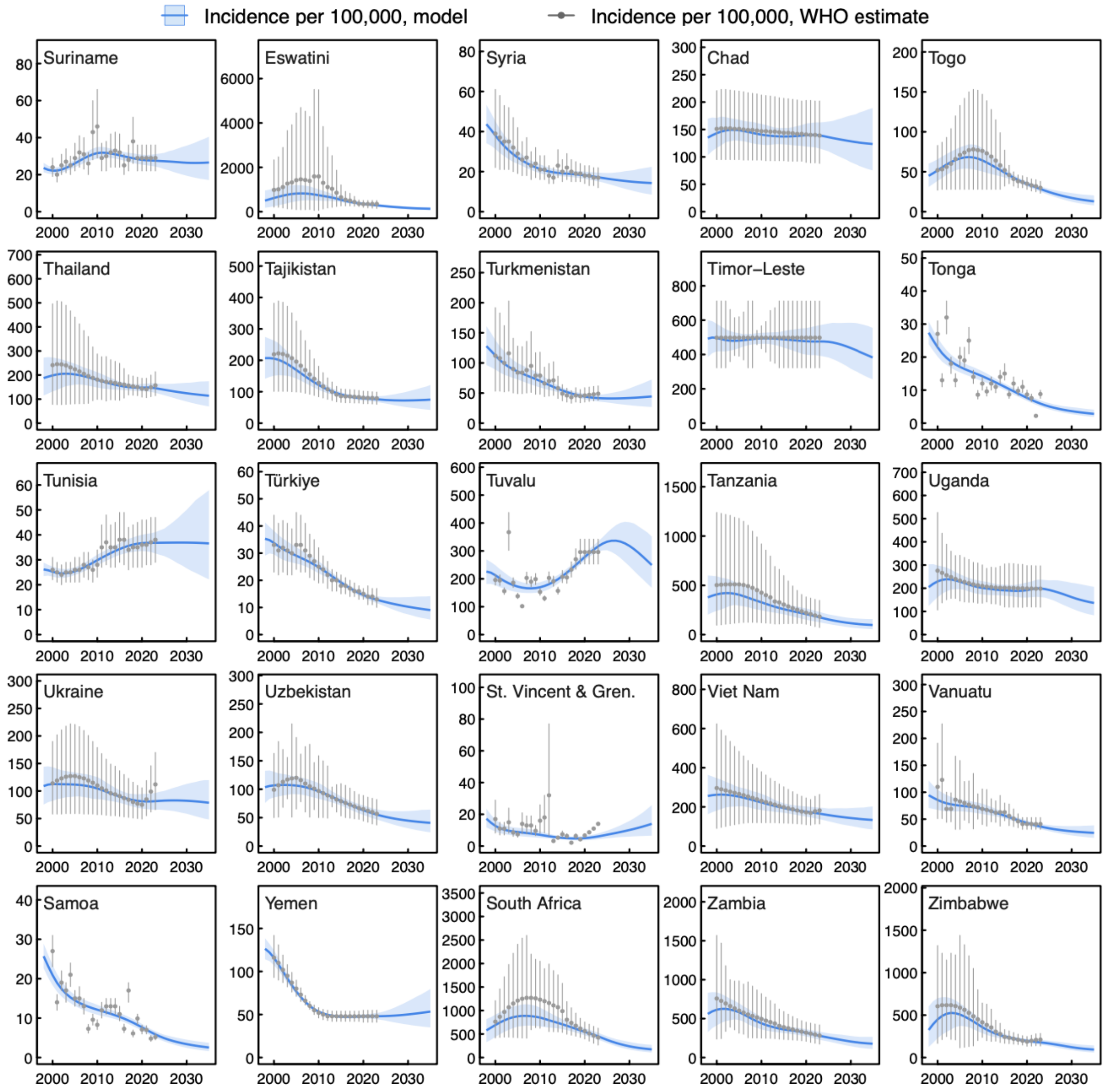

**Figure S2 continued. Fit of country-specific transmission-dynamic models to WHO TB incidence estimates for 2000-2023, with model base-case projections to 2035.**

For WHO estimates, bars represent published uncertainty intervals. For modeled values, shaded region represents 95% uncertainty interval.

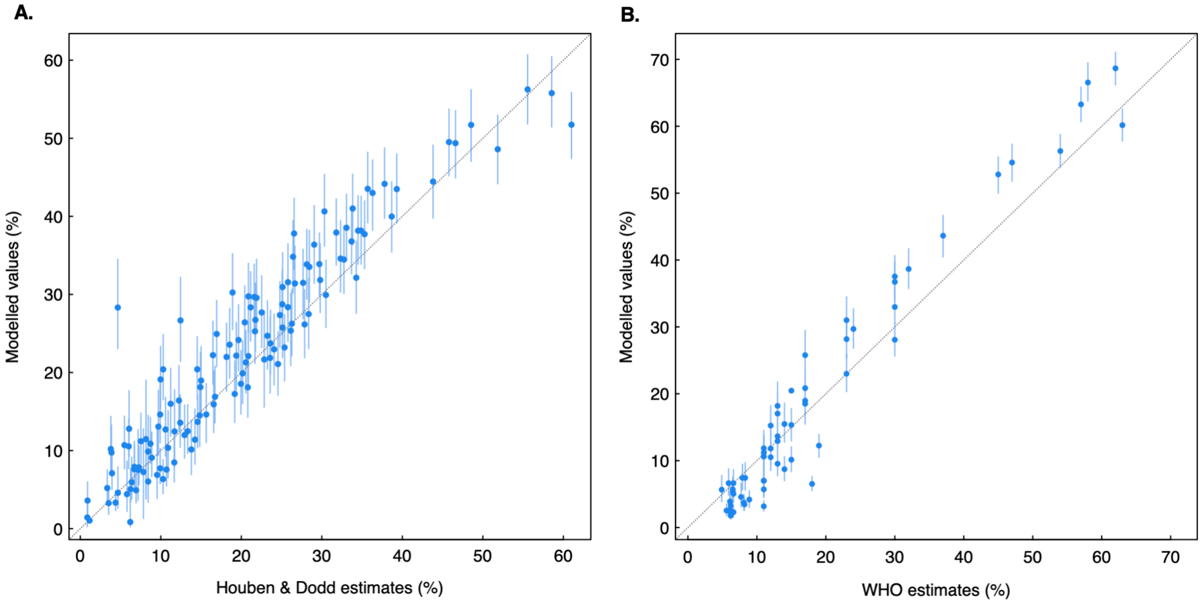

**Figure S2. Fit of country-specific transmission-dynamic models to published estimates of *Mtb* infection prevalence (Panel A) and HIV prevalence among TB cases (Panel B).**

*Mtb* = *Mycobacterium tuberculosis*. *Mtb* infection prevalence compared for 2015 (year available in data source).[17] HIV prevalence among TB cases compared to 2023 WHO estimates[7] for the subset of 63 countries with HIV prevalence >1% or HIV prevalence among TB cases >5%. Bars represent 95% uncertainty intervals.

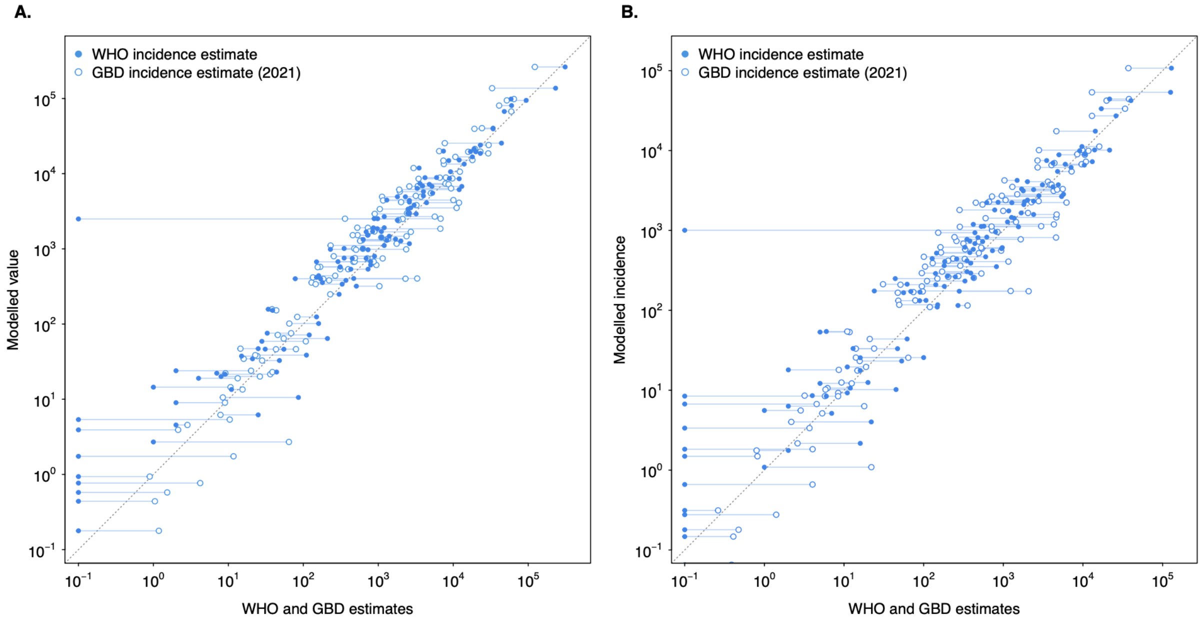

**Figure S3. Modelled estimates of pediatric TB cases in each country for 0-14-year-olds (Panel A) and 0-4-year-olds (Panel B) in 2023, compared to most recent estimates by WHO and the Global Burden of Disease Study.**

WHO estimates[7] equal to zero are plotted at $10^{-1}$. GBD = Global Burden of Disease Study (most recent available year = 2021).[10] Diagonal lines represent lines of equality.

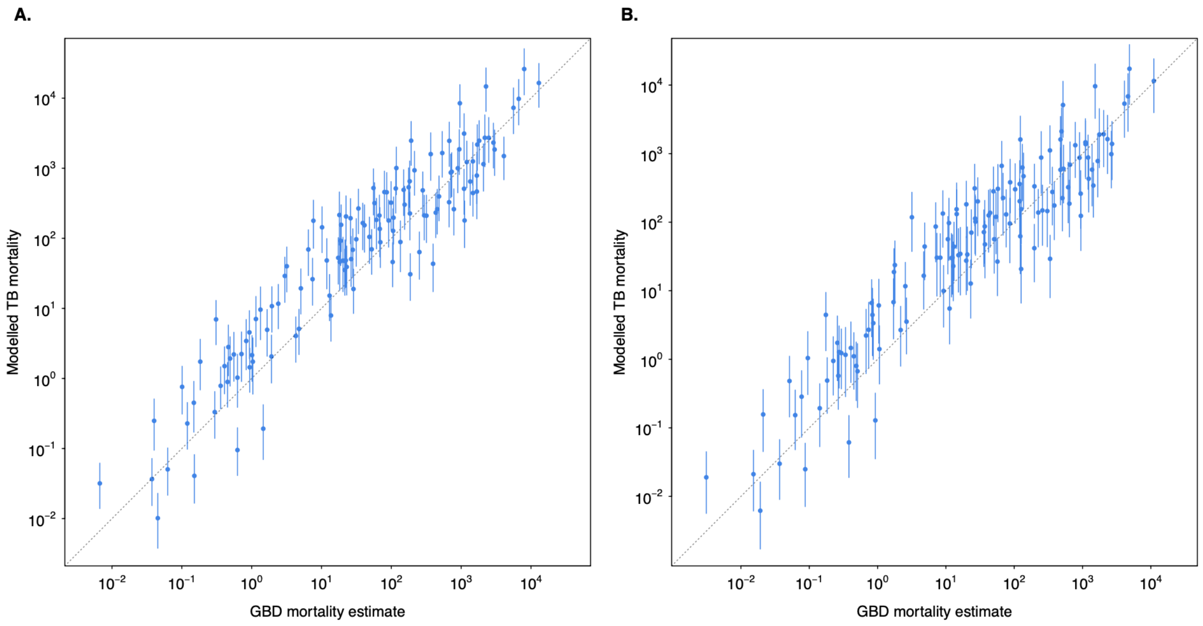

**Figure S4. Modelled estimates of pediatric TB deaths in each country for 0-14-year-olds (Panel A) and 0-4-year-olds (Panel B) in 2023, compared to most recent Global Burden of Disease Study estimates.**

GBD = Global Burden of Disease Study.[10] Diagonal lines represent lines of equality. Vertical bars represent 95% uncertainty intervals.

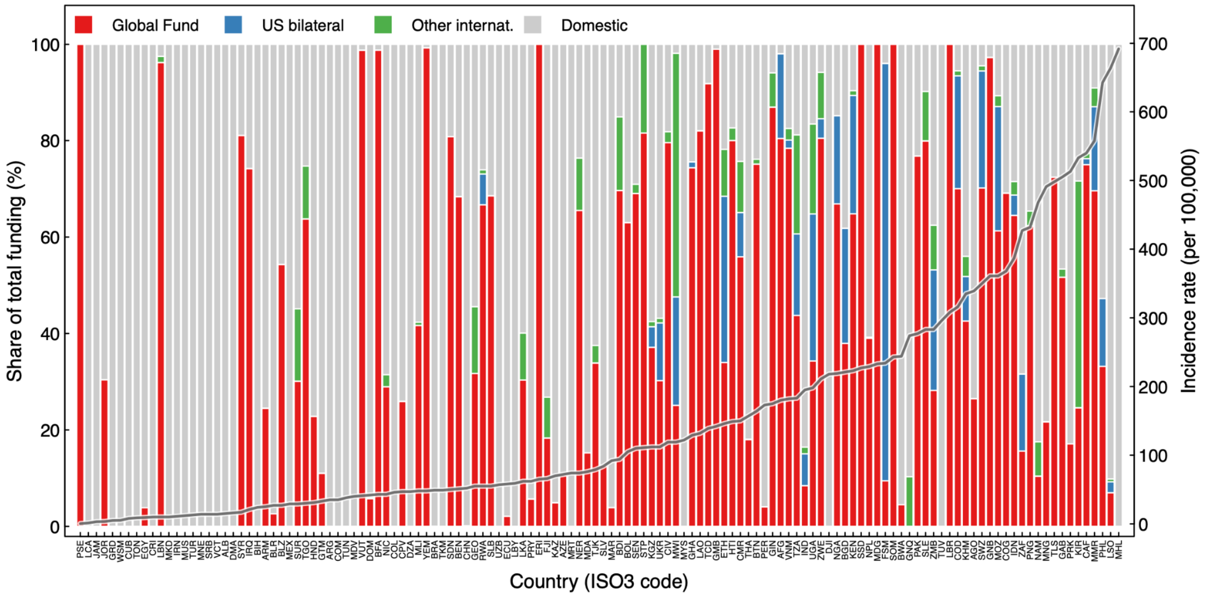

**Figure S5. Contribution of different funding sources to current TB funding in each country prior to 2025.**

Countries ordered by WHO TB incidence rate in 2023.[7]

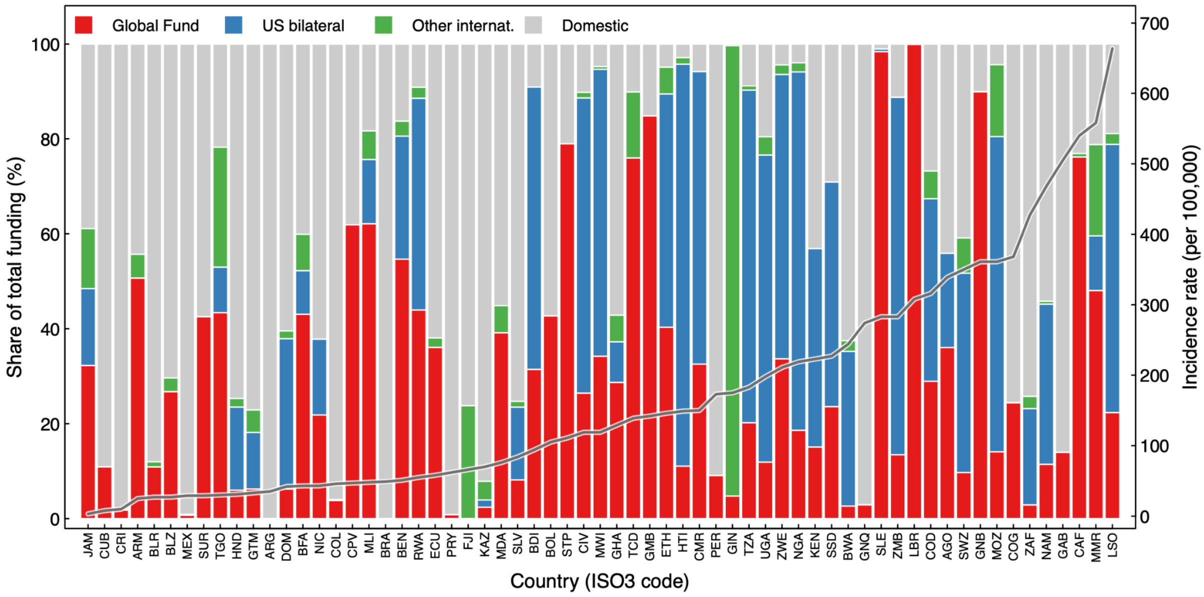

**Figure S6. Contribution of different funding sources to current HIV funding in each country prior to 2025.**

Countries ordered by WHO TB incidence rate in 2023. Results shown for countries with >1% HIV prevalence in general population or HIV prevalence >5% among individuals with TB.[7]

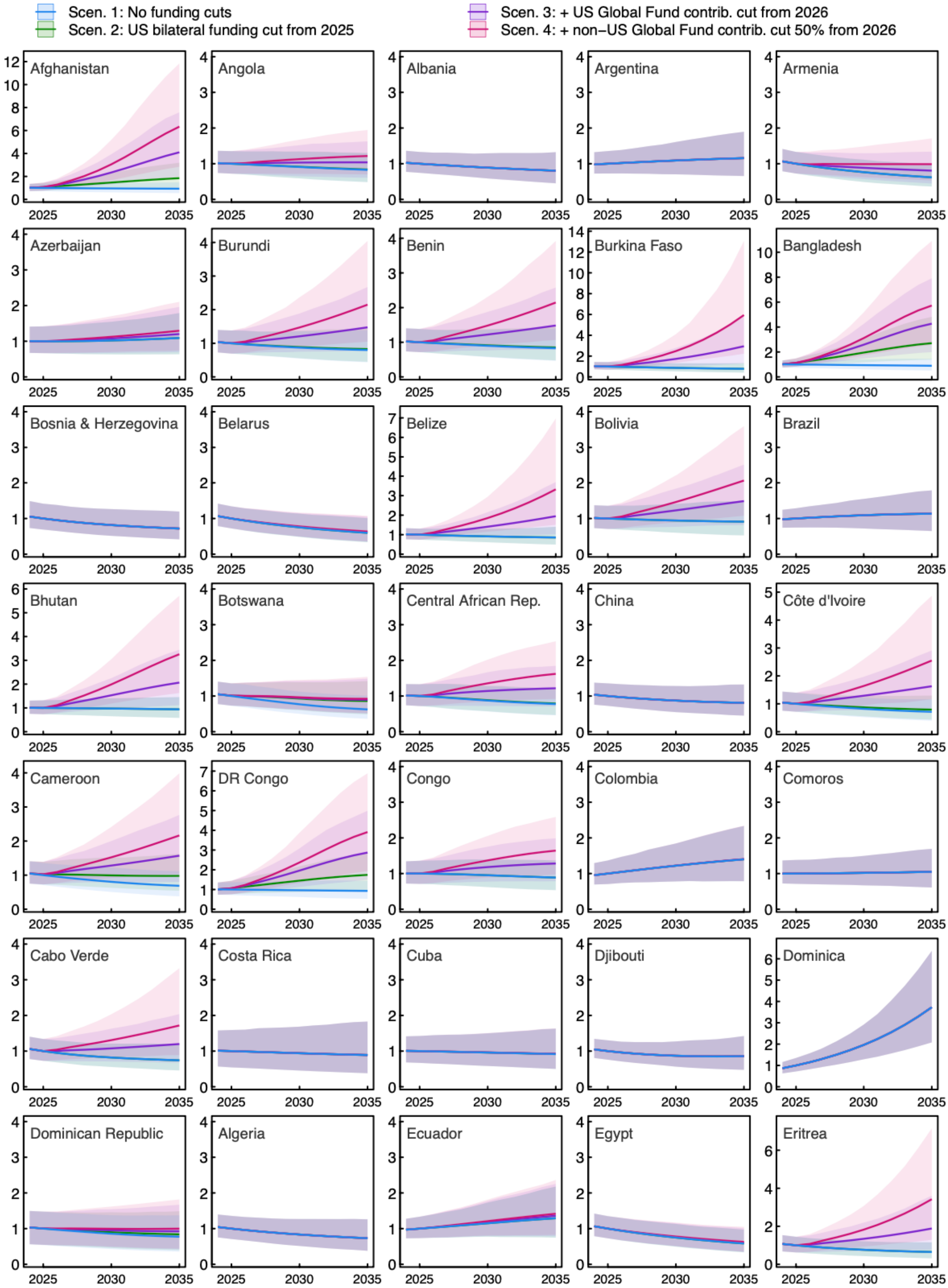

**Figure S7. Change in TB force of infection 2025-2034 relative to the 2024 level in each country, by scenario.**

Shaded regions represent 95% uncertainty intervals. Scen = scenario.

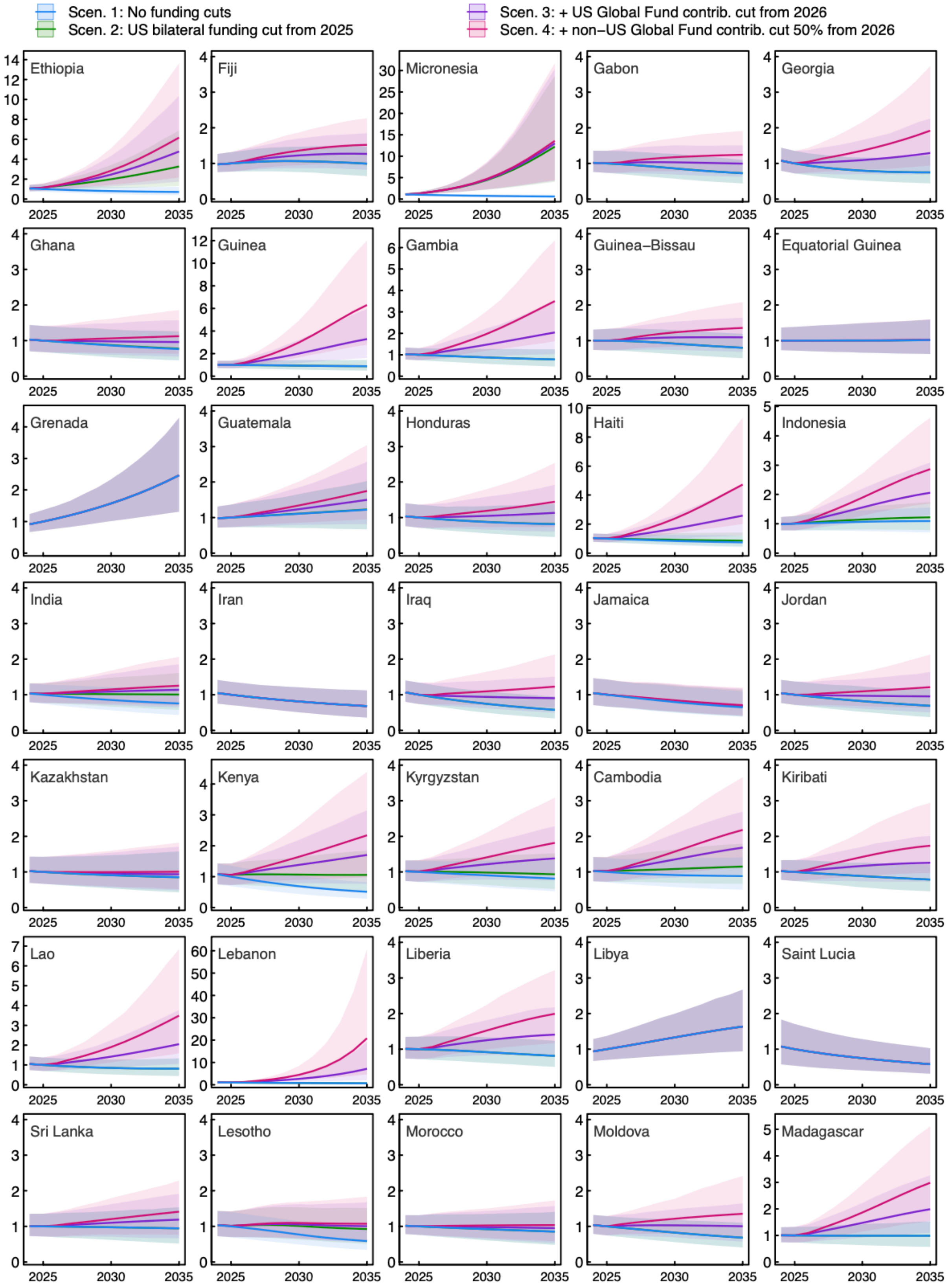

**Figure S7 continued. Change in TB force of infection 2025-2034 relative to the 2024 level in each country, by scenario.**

Shaded regions represent 95% uncertainty intervals. Scen = scenario.

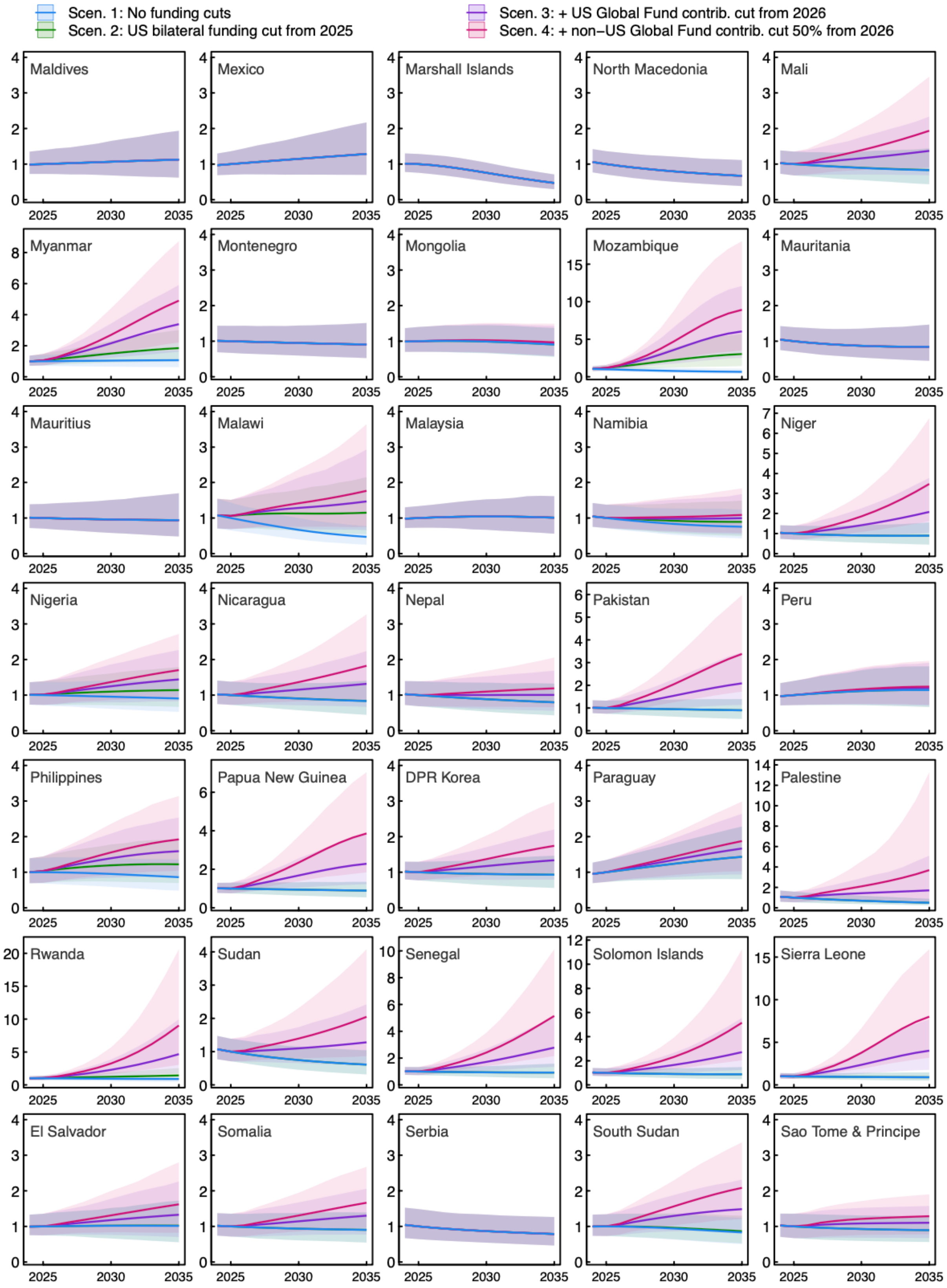

**Figure S7 continued. Change in TB force of infection 2025-2034 relative to the 2024 level in each country, by scenario.**

Shaded regions represent 95% uncertainty intervals. Scen = scenario.

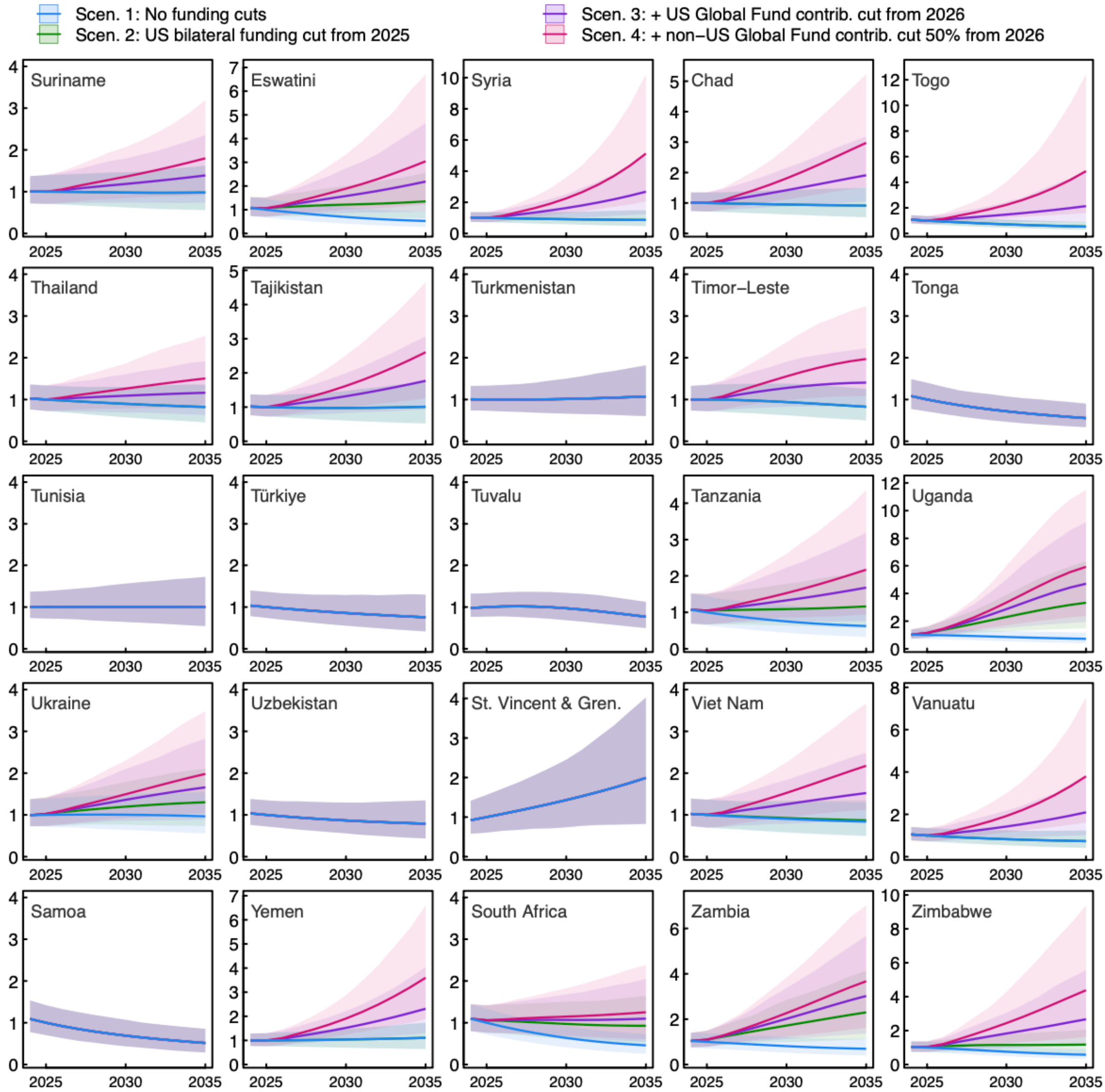

**Figure S7 continued. Change in TB force of infection 2025-2034 relative to the 2024 level in each country, by scenario.**

Shaded regions represent 95% uncertainty intervals. Scen = scenario.

| Country | Reduction in pediatric TB treatment access (percentage points) | | |
|---|---|---|---|
| | Scenario 2 | Scenario 3 | Scenario 4 |
| Micronesia | 35 (20, 54) | 36 (20, 56) | 37 (21, 58) |
| Mozambique | 11 (6, 17) | 21 (12, 32) | 29 (16, 45) |
| Palestine | 0 (0, 0) | 15 (9, 24) | 28 (16, 43) |
| Afghanistan | 6.5 (3.7, 10.1) | 18 (10, 28) | 27 (15, 42) |
| Lebanon | 0 (0, 0) | 15 (8, 23) | 27 (15, 42) |
| Burkina Faso | 0 (0, 0) | 14 (8, 21) | 25 (14, 39) |
| Dem. Rep. Congo | 7.8 (4.4, 12.1) | 17 (9, 26) | 24 (13, 37) |
| Guinea | 0 (0, 0) | 13 (7, 20) | 24 (13, 37) |
| Eritrea | 0 (0, 0) | 13 (7, 20) | 23 (13, 37) |
| Vanuatu | 0 (0, 0) | 13 (7, 20) | 23 (13, 36) |
| Myanmar | 6.1 (3.4, 9.5) | 15 (9, 24) | 23 (13, 36) |
| Sierra Leone | 0 (0, 0) | 13 (7, 20) | 23 (13, 36) |
| Gambia | 0 (0, 0) | 12 (7, 19) | 22 (13, 35) |
| Ethiopia | 13 (7, 21) | 18 (10, 29) | 22 (13, 35) |
| Kenya | 7.4 (4.2, 11.6) | 15 (8, 23) | 21 (12, 33) |
| Madagascar | 0 (0, 0) | 12 (6, 18) | 21 (12, 33) |
| Haiti | 0 (0, 0) | 11 (6, 18) | 21 (12, 32) |
| Yemen | 0 (0, 0) | 11 (6, 18) | 21 (12, 32) |
| Syria | 0 (0, 0) | 11 (6, 18) | 20 (11, 32) |
| Eswatini | 6.8 (3.8, 10.6) | 14 (8, 22) | 20 (11, 32) |
| Rwanda | 2.5 (1.4, 3.8) | 12 (7, 19) | 20 (11, 31) |
| Bangladesh | 9.5 (5.4, 14.8) | 15 (9, 24) | 20 (11, 31) |
| Uganda | 11 (6, 17) | 16 (9, 25) | 20 (11, 31) |
| Lao | 0 (0, 0) | 11 (6, 17) | 20 (11, 30) |
| Chad | 0 (0, 0) | 11 (6, 17) | 19 (11, 30) |
| Zimbabwe | 1.3 (0.7, 2.0) | 11 (6, 17) | 19 (11, 30) |
| Solomon Islands | 0 (0, 0) | 10 (6, 16) | 19 (11, 30) |
| Bhutan | 0 (0, 0) | 10 (6, 16) | 19 (11, 29) |
| South Sudan | 0 (0, 0) | 10 (6, 16) | 19 (10, 29) |
| Pakistan | 0 (0, 0) | 10 (6, 16) | 19 (10, 29) |
| Senegal | 0 (0, 0) | 10 (6, 16) | 18 (10, 28) |
| Liberia | 0 (0, 0) | 9.9 (5.6, 15.5) | 18 (10, 28) |
| Sudan | 0 (0, 0) | 9.8 (5.5, 15.3) | 18 (10, 28) |

| Country | Reduction in pediatric TB treatment access (percentage points) | | |
|---|---|---|---|
| | Scenario 2 | Scenario 3 | Scenario 4 |
| Cote d'Ivoire | 0 (0, 0) | 9.7 (5.5, 15.1) | 18 (10, 27) |
| Togo | 0 (0, 0) | 9.7 (5.4, 15.1) | 17 (10, 27) |
| Viet Nam | 0.50 (0.28, 0.79) | 9.4 (5.3, 14.7) | 17 (9, 26) |
| Papua New Guinea | 0 (0, 0) | 9.1 (5.1, 14.2) | 16 (9, 26) |
| Niger | 0 (0, 0) | 9.1 (5.1, 14.1) | 16 (9, 25) |
| Indonesia | 1.4 (0.8, 2.1) | 9.3 (5.2, 14.5) | 16 (9, 24) |
| Timor-Leste | 0 (0, 0) | 8.3 (4.7, 12.9) | 15 (8, 23) |
| Tanzania | 5.3 (3.0, 8.3) | 11 (6, 16) | 15 (8, 23) |
| Cameroon | 2.8 (1.6, 4.4) | 9.4 (5.3, 14.7) | 15 (8, 23) |
| Zambia | 8.2 (4.6, 12.8) | 12 (7, 18) | 15 (8, 23) |
| Burundi | 0 (0, 0) | 8.1 (4.5, 12.6) | 15 (8, 23) |
| Somalia | 0 (0, 0) | 8.1 (4.5, 12.6) | 15 (8, 23) |
| Benin | 0 (0, 0) | 7.9 (4.4, 12.2) | 14 (8, 22) |
| Belize | 0 (0, 0) | 7.8 (4.4, 12.2) | 14 (8, 22) |
| Iraq | 0 (0, 0) | 7.7 (4.4, 12.1) | 14 (8, 22) |
| Central African Rep. | 0.32 (0.18, 0.51) | 7.8 (4.4, 12.1) | 14 (8, 22) |
| Nigeria | 3.9 (2.2, 6.0) | 9.3 (5.2, 14.5) | 14 (8, 21) |
| Sao Tome & Principe | 0 (0, 0) | 7.3 (4.1, 11.3) | 13 (7, 20) |
| Bolivia | 0 (0, 0) | 7.2 (4.1, 11.3) | 13 (7, 20) |
| Cambodia | 3.1 (1.7, 4.8) | 8.6 (4.8, 13.4) | 13 (7, 20) |
| Congo | 0 (0, 0) | 6.8 (3.9, 10.7) | 12 (7, 19) |
| Philippines | 4.4 (2.5, 6.9) | 8.5 (4.8, 13.2) | 12 (7, 18) |
| Malawi | 6.5 (3.6, 10.1) | 9.2 (5.2, 14.4) | 11 (6, 18) |
| Guinea-Bissau | 0 (0, 0) | 6.3 (3.5, 9.8) | 11 (6, 18) |
| Kyrgyzstan | 1.4 (0.8, 2.2) | 6.2 (3.5, 9.7) | 10 (6, 16) |
| Ukraine | 3.7 (2.1, 5.7) | 7.2 (4.0, 11.2) | 10 (6, 16) |
| Mali | 0 (0, 0) | 5.4 (3.0, 8.4) | 9.8 (5.5, 15.3) |
| Gabon | 0 (0, 0) | 5.2 (2.9, 8.1) | 9.4 (5.3, 14.6) |
| Ghana | 0.21 (0.12, 0.33) | 5.2 (3.0, 8.2) | 9.3 (5.3, 14.5) |
| Tajikistan | 0.012 (0.007, 0.019) | 4.9 (2.8, 7.6) | 8.8 (5.0, 13.8) |
| Georgia | 0 (0, 0) | 4.8 (2.7, 7.5) | 8.7 (4.9, 13.6) |
| South Africa | 5.2 (2.9, 8.1) | 7.1 (4.0, 11.1) | 8.7 (4.9, 13.5) |
| Nicaragua | 0 (0, 0) | 4.1 (2.3, 6.4) | 7.4 (4.2, 11.5) |
| Cabo Verde | 0 (0, 0) | 4.0 (2.2, 6.2) | 7.2 (4.0, 11.2) |

| Country | Reduction in pediatric TB treatment access (percentage points) | | |
|---|---|---|---|
| | Scenario 2 | Scenario 3 | Scenario 4 |
| Suriname | 0 (0, 0) | 3.9 (2.2, 6.1) | 7.1 (4.0, 11.1) |
| Nepal | 0 (0, 0) | 3.8 (2.1, 5.9) | 6.8 (3.9, 10.7) |
| Jordan | 0 (0, 0) | 3.8 (2.1, 5.9) | 6.8 (3.8, 10.6) |
| Kiribati | 0 (0, 0) | 3.8 (2.1, 5.9) | 6.8 (3.8, 10.6) |
| Sri Lanka | 0 (0, 0) | 3.3 (1.9, 5.2) | 6.0 (3.4, 9.4) |
| Honduras | 0 (0, 0) | 3.2 (1.8, 5.0) | 5.8 (3.2, 9.0) |
| Armenia | 0 (0, 0) | 3.1 (1.8, 4.9) | 5.7 (3.2, 8.8) |
| Angola | 0 (0, 0) | 3.0 (1.7, 4.6) | 5.3 (3.0, 8.3) |
| Thailand | 0 (0, 0) | 2.7 (1.5, 4.2) | 4.9 (2.8, 7.7) |
| DPR Korea | 0 (0, 0) | 2.6 (1.5, 4.1) | 4.8 (2.7, 7.4) |
| Fiji | 0 (0, 0) | 2.5 (1.4, 3.9) | 4.5 (2.6, 7.1) |
| India | 2.4 (1.4, 3.8) | 3.6 (2.0, 5.6) | 4.5 (2.5, 7.1) |
| Moldova | 0 (0, 0) | 2.3 (1.3, 3.6) | 4.2 (2.4, 6.5) |
| El Salvador | 0 (0, 0) | 1.9 (1.1, 3.0) | 3.5 (2.0, 5.4) |
| Guatemala | 0 (0, 0) | 1.6 (0.9, 2.5) | 2.9 (1.6, 4.5) |
| Azerbaijan | 0 (0, 0) | 1.4 (0.8, 2.1) | 2.4 (1.4, 3.8) |
| Namibia | 0 (0, 0) | 1.2 (0.7, 1.8) | 2.1 (1.2, 3.3) |
| Mongolia | 0 (0, 0) | 1.1 (0.6, 1.7) | 1.9 (1.1, 3.0) |
| Paraguay | 0 (0, 0) | 0.87 (0.49, 1.35) | 1.6 (0.9, 2.4) |
| Dominican Republic | 0 (0, 0) | 0.83 (0.47, 1.30) | 1.5 (0.8, 2.3) |
| Lesotho | 0.46 (0.26, 0.72) | 1.0 (0.6, 1.6) | 1.5 (0.8, 2.3) |
| Kazakhstan | 0 (0, 0) | 0.69 (0.39, 1.08) | 1.3 (0.7, 2.0) |
| Morocco | 0 (0, 0) | 0.59 (0.34, 0.93) | 1.1 (0.6, 1.7) |
| Peru | 0 (0, 0) | 0.52 (0.29, 0.81) | 0.94 (0.53, 1.46) |
| Egypt | 0 (0, 0) | 0.47 (0.26, 0.73) | 0.84 (0.48, 1.32) |
| Botswana | 0 (0, 0) | 0.46 (0.26, 0.71) | 0.83 (0.47, 1.29) |
| Belarus | 0 (0, 0) | 0.35 (0.20, 0.54) | 0.63 (0.36, 0.98) |
| Ecuador | 0 (0, 0) | 0.31 (0.17, 0.48) | 0.56 (0.31, 0.87) |

**Table S2. Assumed reduction in pediatric TB treatment access by 2026 under each funding reduction scenario, as compared to the base-case scenario.**

Results presented in order of decreasing reduction in coverage under Scenario 4 (final column). Values in parentheses represent 95% uncertainty intervals. Scenario 2: 100% of US bilateral funding cut in 2025. Scenario 3: 100% of US bilateral funding cut in 2025, plus 100% of US Global Fund contributions cut in 2026. Scenario 4: 100% of US bilateral funding cut in 2025, plus 100% of US and 50% of non-US Global Fund contributions cut in 2026.

| | Pediatric TB cases | | | Pediatric TB deaths | | |
|---|---|---|---|---|---|---|
| | 2025-2029 | 2030-2034 | Whole period | 2025-2029 | 2030-2034 | Whole period |
| *Absolute increase (millions)* | | | | | | |
| Scen. 2: US bilateral funding cut in 2025 | 0.62 (0.47, 0.81) | 1.85 (1.34, 2.52) | 2.47 (1.81, 3.34) | 0.10 (0.08, 0.14) | 0.23 (0.17, 0.32) | 0.34 (0.24, 0.46) |
| Scen. 3: + US Global Fund contributions cut in 2026 | 1.22 (0.96, 1.53) | 4.67 (3.61, 6.10) | 5.89 (4.58, 7.60) | 0.24 (0.18, 0.31) | 0.68 (0.51, 0.93) | 0.92 (0.70, 1.23) |
| Scen. 4: + 50% of non-US Global Fund contributions cut in 2026 | 1.65 (1.31, 2.07) | 7.23 (5.57, 9.42) | 8.89 (6.88, 11.45) | 0.36 (0.28, 0.46) | 1.17 (0.86, 1.59) | 1.52 (1.14, 2.03) |
| *Relative increase (%)* | | | | | | |
| Scen. 2: US bilateral funding cut in 2025 | 11 (8, 13) | 34 (26, 45) | 22 (17, 28) | 18 (14, 23) | 44 (33, 59) | 31 (24, 40) |
| Scen. 3: + US Global Fund contributions cut in 2026 | 21 (17, 25) | 86 (69, 107) | 52 (43, 64) | 42 (35, 51) | 128 (102, 166) | 84 (67, 106) |
| Scen. 4: + 50% of non-US Global Fund contributions cut in 2026 | 28 (24, 34) | 133 (108, 165) | 79 (64, 96) | 63 (52, 76) | 219 (172, 288) | 139 (110, 178) |

**Table S3: Additional pediatric TB cases and deaths in low- and middle-income countries projected under alternative funding scenarios as compared to continued funding, 2025-2034.**

Scen = scenario. Scenarios 2-4 represent progressively greater funding cuts, such that Scenario 4 includes 100% reductions in US bilateral funding in 2025 and Global Fund support in 2026, and 50% reductions in non-US Global Fund support in 2026.

| | Thousands of pediatric TB cases | | | Thousands of pediatric TB deaths | | |
|---|---|---|---|---|---|---|
| | Scenario 2 | Scenario 3 | Scenario 4 | Scenario 2 | Scenario 3 | Scenario 4 |
| Total | 22<br>(17, 28) | 52<br>(43, 64) | 79<br>(64, 96) | 31<br>(24, 40) | 84<br>(67, 106) | 139<br>(110, 178) |
| WHO region | | | | | | |
| *Eastern Mediterranean Region* | 5.2<br>(1.6, 12.5) | 55<br>(36, 84) | 101<br>(64, 155) | 6.7<br>(1.9, 17.2) | 90<br>(56, 144) | 185<br>(109, 301) |
| *African Region* | 36<br>(25, 51) | 74<br>(55, 102) | 108<br>(81, 146) | 44<br>(30, 64) | 104<br>(74, 150) | 165<br>(118, 241) |
| *European Region* | 3.3<br>(1.5, 6.2) | 14<br>(9, 21) | 23<br>(15, 35) | 6.0<br>(2.6, 11.0) | 23<br>(14, 34) | 39<br>(24, 59) |
| *Region of the Americas* | 0.45<br>(0.18, 0.92) | 7.0<br>(3.6, 13.0) | 13<br>(7, 25) | 0.52<br>(0.21, 1.12) | 14<br>(7, 26) | 30<br>(14, 57) |
| *South-East Asian Region* | 20<br>(12, 30) | 43<br>(29, 62) | 63<br>(42, 91) | 28<br>(17, 43) | 71<br>(46, 106) | 114<br>(73, 173) |
| *Western Pacific Region* | 12<br>(6, 19) | 29<br>(18, 42) | 44<br>(28, 63) | 21<br>(11, 36) | 56<br>(35, 85) | 90<br>(58, 133) |
| Income group | | | | | | |
| *Low income* | 49<br>(33, 73) | 105<br>(75, 145) | 154<br>(113, 209) | 65<br>(41, 98) | 163<br>(112, 236) | 267<br>(183, 388) |
| *Lower middle income* | 18<br>(13, 25) | 43<br>(33, 56) | 65<br>(49, 86) | 25<br>(18, 34) | 66<br>(50, 87) | 109<br>(79, 149) |
| *Upper middle income* | 5.7<br>(3.4, 8.9) | 27<br>(15, 43) | 44<br>(25, 73) | 9.9<br>(6.0, 15.4) | 52<br>(29, 86) | 94<br>(49, 163) |
| TB burden category* | | | | | | |
| *High TB burden* | 25<br>(19, 32) | 54<br>(43, 69) | 80<br>(63, 101) | 35<br>(26, 47) | 88<br>(68, 115) | 142<br>(108, 187) |
| *Not high TB burden* | 7.3<br>(4.5, 11.5) | 41<br>(32, 53) | 72<br>(57, 90) | 9.0<br>(5.6, 15.1) | 64<br>(51, 85) | 125<br>(98, 166) |
| Funding category** | | | | | | |
| *High non-domestic funding* | 38<br>(27, 53) | 82<br>(62, 109) | 120<br>(92, 159) | 47<br>(33, 68) | 117<br>(86, 166) | 190<br>(137, 270) |
| *Non high non-domestic funding* | 13<br>(8, 19) | 36<br>(26, 47) | 56<br>(40, 75) | 18<br>(12, 28) | 59<br>(43, 81) | 101<br>(71, 144) |

**Table S4: Percentage increase in pediatric TB cases and deaths under alternative funding scenarios as compared to continued funding, 2025-2034, by country grouping.**

Scenario 2: 100% of US bilateral funding cut in 2025. Scenario 3: 100% of US bilateral funding cut in 2025, plus 100% of US Global Fund contributions cut in 2026. Scenario 4: 100% of US bilateral funding cut in 2025, plus 100% of US and 50% of non-US Global Fund contributions cut in 2026. *High TB burden group includes the 30 countries identified as having high TB burden by WHO. ** High non-domestic funding group represents countries with >80% non-domestic funding for TB or HIV programs.

| | Thousands of pediatric TB cases | | | Thousands of pediatric TB deaths | | |
|---|---|---|---|---|---|---|
| | Scenario 2 | Scenario 3 | Scenario 4 | Scenario 2 | Scenario 3 | Scenario 4 |
| Dem. Rep. Congo | 334 (112, 719) | 697 (227, 1,496) | 1,003 (326, 2,169) | 54 (17, 125) | 128 (38, 299) | 202 (58, 479) |
| Indonesia | 88 (32, 190) | 550 (196, 1,195) | 945 (333, 2,057) | 13 (5, 29) | 93 (32, 210) | 173 (59, 399) |
| Pakistan | 0 (0, 0) | 482 (161, 1,109) | 927 (302, 2,135) | 0 (0, 0) | 75 (22, 186) | 161 (45, 419) |
| Bangladesh | 324 (114, 733) | 539 (185, 1,234) | 726 (246, 1,673) | 35 (11, 85) | 65 (19, 161) | 95 (27, 238) |
| Mozambique | 247 (80, 610) | 491 (155, 1,211) | 705 (221, 1,722) | 25 (7, 63) | 62 (16, 161) | 102 (25, 273) |
| India | 313 (98, 662) | 448 (138, 953) | 561 (172, 1,197) | 37 (11, 79) | 54 (17, 117) | 69 (21, 150) |
| Ethiopia | 251 (81, 598) | 360 (111, 881) | 454 (139, 1,120) | 32 (9, 85) | 50 (13, 133) | 67 (16, 182) |
| Myanmar | 97 (33, 210) | 241 (77, 539) | 366 (119, 803) | 13 (4, 28) | 37 (11, 84) | 62 (17, 144) |
| Uganda | 204 (60, 495) | 290 (84, 698) | 362 (105, 877) | 24 (7, 64) | 38 (10, 104) | 51 (13, 143) |
| Nigeria | 124 (45, 266) | 253 (92, 542) | 359 (131, 765) | 30 (10, 69) | 67 (23, 154) | 99 (33, 228) |
| Philippines | 143 (52, 308) | 258 (93, 561) | 354 (127, 772) | 22 (8, 49) | 43 (15, 95) | 61 (21, 138) |
| Afghanistan | 65 (22, 147) | 185 (60, 423) | 289 (93, 668) | 8.2 (2.6, 20.8) | 28 (8, 72) | 50 (14, 130) |
| Tanzania | 56 (18, 134) | 98 (30, 236) | 133 (41, 324) | 8.7 (2.7, 22.4) | 17 (5, 43) | 24 (7, 64) |
| Madagascar | 0 (0, 0) | 68 (25, 149) | 127 (47, 276) | 0 (0, 0) | 14 (4, 32) | 28 (9, 67) |
| Kenya | 46 (15, 103) | 85 (28, 190) | 119 (39, 266) | 7.8 (2.4, 18.1) | 16 (5, 38) | 24 (7, 59) |
| Zambia | 68 (21, 158) | 92 (27, 219) | 112 (33, 270) | 9.3 (2.7, 22.5) | 13 (4, 33) | 17 (5, 42) |
| Guinea | 0 (0, 0) | 50 (16, 121) | 102 (33, 244) | 0 (0, 0) | 6.3 (1.7, 16.5) | 15 (4, 41) |
| Sierra Leone | 0.012 (0.004, 0.028) | 48 (15, 121) | 100 (31, 251) | 0.001 (0.000, 0.001) | 4.9 (1.4, 12.4) | 13 (3, 33) |
| Viet Nam | 3.2 (1.1, 6.8) | 52 (18, 112) | 94 (32, 201) | 0.52 (0.18, 1.09) | 9.5 (3.1, 20.6) | 19 (6, 40) |
| Papua New Guinea | 0 (0, 0) | 38 (13, 85) | 75 (25, 172) | 0 (0, 0) | 4.5 (1.3, 10.6) | 10 (3, 24) |
| South Africa | 42 (14, 92) | 54 (17, 119) | 64 (20, 142) | 6.0 (1.9, 13.8) | 7.9 (2.4, 18.4) | 9.6 (2.9, 22.5) |
| Chad | 0 (0, 0) | 31 (11, 70) | 58 (20, 135) | 0 (0, 0) | 6.2 (1.9, 15.0) | 13 (4, 31) |
| Zimbabwe | 13 (4, 29) | 35 (10, 81) | 57 (16, 131) | 1.4 (0.4, 3.3) | 5.3 (1.4, 12.6) | 9.8 (2.4, 24.0) |
| Cote d'Ivoire | 3.1 (1.0, 7.3) | 29 (10, 64) | 52 (17, 118) | 0.33 (0.10, 0.76) | 4.9 (1.5, 11.9) | 9.9 (2.8, 25.1) |
| Cameroon | 12 (4, 27) | 30 (10, 70) | 46 (15, 107) | 1.8 (0.6, 4.2) | 5.2 (1.7, 12.6) | 8.6 (2.7, 21.4) |
| Angola | 1.3 (0.4, 3.1) | 25 (9, 55) | 45 (16, 98) | 0.17 (0.05, 0.40) | 4.6 (1.5, 10.8) | 8.5 (2.8, 19.9) |

| | Thousands of pediatric TB cases | | | Thousands of pediatric TB deaths | | |
|---|---|---|---|---|---|---|
| | Scenario 2 | Scenario 3 | Scenario 4 | Scenario 2 | Scenario 3 | Scenario 4 |
| Cambodia | 11 (4, 24) | 29 (10, 63) | 44 (15, 96) | 1.6 (0.5, 3.6) | 4.5 (1.4, 10.4) | 7.3 (2.3, 17.3) |
| Senegal | 0 (0, 0) | 22 (7, 51) | 43 (14, 105) | 0 (0, 0) | 2.6 (0.8, 6.4) | 6.0 (1.7, 15.0) |
| Niger | 0 (0, 0) | 22 (7, 49) | 42 (14, 97) | 0 (0, 0) | 3.2 (1.0, 8.0) | 6.9 (2.0, 17.9) |
| Yemen | 0 (0, 0) | 22 (8, 47) | 42 (15, 90) | 0 (0, 0) | 4.4 (1.5, 11.1) | 9.2 (3.0, 23.5) |
| Somalia | 0 (0, 0) | 22 (8, 47) | 40 (14, 86) | 0 (0, 0) | 6.0 (2.0, 14.3) | 12 (4, 28) |
| Central African Rep. | 0.91 (0.35, 1.87) | 22 (8, 46) | 39 (15, 84) | 0.20 (0.07, 0.43) | 5.0 (1.7, 11.4) | 9.7 (3.3, 22.4) |
| Haiti | 1.4 (0.5, 3.1) | 16 (6, 37) | 31 (10, 70) | 0.11 (0.04, 0.24) | 2.0 (0.6, 4.9) | 4.5 (1.4, 11.4) |
| South Sudan | 0.97 (0.34, 2.15) | 17 (6, 35) | 30 (11, 62) | 0.14 (0.05, 0.32) | 4.0 (1.4, 8.5) | 7.8 (2.6, 16.9) |
| Burkina Faso | 0.038 (0.010, 0.091) | 14 (4, 34) | 28 (9, 70) | 0.003 (0.001, 0.008) | 2.1 (0.6, 5.2) | 4.8 (1.3, 12.6) |
| Sudan | 0 (0, 0) | 14 (5, 31) | 27 (9, 60) | 0 (0, 0) | 2.6 (0.8, 6.2) | 5.4 (1.6, 13.0) |
| Rwanda | 2.7 (0.8, 5.9) | 13 (4, 31) | 24 (7, 58) | 0.23 (0.07, 0.54) | 1.4 (0.4, 3.7) | 3.1 (0.8, 8.2) |
| DPR Korea | 0 (0, 0) | 11 (4, 26) | 21 (7, 50) | 0 (0, 0) | 0.99 (0.33, 2.27) | 2.0 (0.6, 4.6) |
| Malawi | 12 (3, 28) | 16 (5, 38) | 20 (5, 48) | 2.0 (0.6, 4.9) | 2.8 (0.8, 7.0) | 3.6 (0.9, 9.1) |
| Liberia | 0 (0, 0) | 9.8 (3.5, 21.5) | 18 (6, 40) | 0 (0, 0) | 2.2 (0.7, 5.1) | 4.4 (1.4, 10.4) |
| Thailand | 0 (0, 0) | 8.7 (2.7, 19.2) | 16 (5, 37) | 0 (0, 0) | 0.82 (0.26, 1.91) | 1.6 (0.5, 3.8) |
| Congo | 0 (0, 0) | 8.6 (3.0, 18.6) | 16 (5, 34) | 0 (0, 0) | 2.0 (0.6, 4.5) | 3.8 (1.2, 8.9) |
| Ghana | 0.62 (0.21, 1.39) | 8.2 (2.8, 17.8) | 14 (5, 31) | 0.15 (0.05, 0.34) | 2.3 (0.8, 5.4) | 4.3 (1.4, 9.8) |
| Burundi | 0.39 (0.12, 0.95) | 7.5 (2.6, 16.9) | 14 (5, 31) | 0.044 (0.014, 0.106) | 1.4 (0.5, 3.2) | 2.7 (0.9, 6.5) |
| Lao | 0 (0, 0) | 7.2 (2.4, 16.8) | 14 (5, 33) | 0 (0, 0) | 1.0 (0.3, 2.6) | 2.3 (0.7, 5.9) |
| Mali | 0.088 (0.026, 0.221) | 6.4 (2.3, 14.4) | 12 (4, 28) | 0.009 (0.003, 0.023) | 0.99 (0.32, 2.34) | 2.0 (0.6, 4.9) |
| Nepal | 0 (0, 0) | 5.7 (1.9, 13.5) | 10 (3, 25) | 0 (0, 0) | 1.2 (0.4, 2.7) | 2.2 (0.7, 5.0) |
| Benin | 0.22 (0.06, 0.53) | 5.0 (1.7, 11.3) | 9.2 (3.0, 20.9) | 0.025 (0.007, 0.060) | 0.93 (0.29, 2.18) | 1.8 (0.5, 4.4) |
| Bolivia | 0 (0, 0) | 4.5 (1.6, 9.9) | 8.3 (2.9, 18.6) | 0 (0, 0) | 0.80 (0.26, 1.82) | 1.6 (0.5, 3.7) |
| Syria | 0 (0, 0) | 3.5 (1.1, 7.8) | 6.9 (2.1, 16.0) | 0 (0, 0) | 0.52 (0.14, 1.31) | 1.2 (0.3, 3.1) |
| Gambia | 0 (0, 0) | 3.4 (1.1, 7.6) | 6.5 (2.2, 15.0) | 0 (0, 0) | 0.58 (0.18, 1.38) | 1.3 (0.4, 3.1) |
| Tajikistan | 0.008 (0.003, 0.019) | 3.0 (0.9, 7.3) | 5.8 (1.8, 14.0) | 0.001 (0.000, 0.002) | 0.34 (0.10, 0.84) | 0.71 (0.20, 1.74) |

| | Thousands of pediatric TB cases | | | Thousands of pediatric TB deaths | | |
|---|---|---|---|---|---|---|
| | Scenario 2 | Scenario 3 | Scenario 4 | Scenario 2 | Scenario 3 | Scenario 4 |
| Togo | 0.020 (0.006, 0.053) | 2.5 (0.7, 6.2) | 5.3 (1.4, 14.0) | 0.001 (0.000, 0.003) | 0.26 (0.07, 0.69) | 0.64 (0.17, 1.78) |
| Gabon | 0 (0, 0) | 2.8 (1.0, 6.2) | 5.2 (1.8, 11.4) | 0 (0, 0) | 0.58 (0.19, 1.29) | 1.1 (0.4, 2.5) |
| Timor-Leste | 0 (0, 0) | 2.6 (0.9, 5.9) | 4.9 (1.6, 10.8) | 0 (0, 0) | 0.51 (0.15, 1.20) | 1.0 (0.3, 2.4) |
| Guinea-Bissau | 0 (0, 0) | 2.5 (0.9, 5.4) | 4.6 (1.7, 9.9) | 0 (0, 0) | 0.70 (0.24, 1.54) | 1.3 (0.5, 3.0) |
| Ukraine | 1.6 (0.5, 3.5) | 2.9 (0.9, 6.4) | 4.1 (1.3, 8.8) | 0.25 (0.08, 0.55) | 0.49 (0.15, 1.10) | 0.70 (0.21, 1.58) |
| Iraq | 0 (0, 0) | 2.0 (0.7, 4.5) | 3.8 (1.3, 8.3) | 0 (0, 0) | 0.45 (0.14, 1.03) | 0.88 (0.27, 2.03) |
| Lesotho | 2.6 (0.8, 6.6) | 3.1 (1.0, 7.8) | 3.5 (1.1, 8.9) | 0.45 (0.14, 1.12) | 0.54 (0.17, 1.37) | 0.63 (0.19, 1.57) |
| Eritrea | 0 (0, 0) | 1.8 (0.6, 4.0) | 3.5 (1.1, 7.9) | 0 (0, 0) | 0.31 (0.09, 0.76) | 0.68 (0.19, 1.74) |
| Sri Lanka | 0 (0, 0) | 1.5 (0.5, 3.4) | 2.7 (0.9, 6.3) | 0 (0, 0) | 0.28 (0.08, 0.67) | 0.52 (0.16, 1.30) |
| Eswatini | 1.1 (0.4, 2.7) | 1.9 (0.6, 4.8) | 2.6 (0.8, 6.5) | 0.18 (0.06, 0.45) | 0.35 (0.10, 0.92) | 0.52 (0.14, 1.39) |
| Peru | 0 (0, 0) | 1.4 (0.5, 3.1) | 2.6 (0.9, 5.6) | 0 (0, 0) | 0.19 (0.06, 0.42) | 0.34 (0.11, 0.77) |
| Morocco | 0 (0, 0) | 1.3 (0.4, 3.1) | 2.3 (0.7, 5.7) | 0 (0, 0) | 0.088 (0.029, 0.204) | 0.16 (0.05, 0.38) |
| Kyrgyzstan | 0.35 (0.11, 0.76) | 1.4 (0.4, 3.0) | 2.3 (0.7, 5.0) | 0.048 (0.015, 0.111) | 0.20 (0.06, 0.49) | 0.35 (0.11, 0.85) |
| Namibia | 0.96 (0.28, 2.34) | 1.6 (0.5, 3.8) | 2.1 (0.6, 5.0) | 0.12 (0.03, 0.28) | 0.22 (0.06, 0.53) | 0.31 (0.09, 0.75) |
| Lebanon | 0 (0, 0) | 0.76 (0.20, 2.00) | 1.8 (0.4, 4.9) | 0 (0, 0) | 0.071 (0.017, 0.200) | 0.21 (0.04, 0.65) |
| Guatemala | 0.025 (0.007, 0.063) | 0.91 (0.32, 2.03) | 1.7 (0.6, 3.7) | 0.002 (0.000, 0.004) | 0.089 (0.028, 0.197) | 0.17 (0.05, 0.38) |
| Nicaragua | 0.007 (0.002, 0.016) | 0.69 (0.22, 1.55) | 1.3 (0.4, 3.0) | 0.000 (0.000, 0.001) | 0.079 (0.024, 0.190) | 0.16 (0.05, 0.38) |
| Bhutan | 0 (0, 0) | 0.63 (0.21, 1.40) | 1.2 (0.4, 2.6) | 0 (0, 0) | 0.091 (0.028, 0.216) | 0.19 (0.06, 0.47) |
| Honduras | 0.011 (0.003, 0.026) | 0.59 (0.20, 1.31) | 1.1 (0.4, 2.5) | 0.001 (0.000, 0.002) | 0.071 (0.024, 0.162) | 0.14 (0.04, 0.32) |
| Solomon Islands | 0 (0, 0) | 0.53 (0.17, 1.22) | 1.1 (0.3, 2.5) | 0 (0, 0) | 0.062 (0.018, 0.147) | 0.14 (0.04, 0.36) |

**Table S5: Additional pediatric TB cases and deaths under alternative funding scenarios as compared to continued funding, 2025-2034, by country.**

Countries ordered by additional TB incidence for Scenario 4. Excludes countries with <1000 additional TB cases projected for Scenario 4. Scenario 2: 100% of US bilateral funding cut in 2025. Scenario 3: 100% of US bilateral funding cut in 2025, plus 100% of US Global Fund contributions cut in 2026. Scenario 4: 100% of US bilateral funding cut in 2025, plus 100% of US and 50% of non-US Global Fund contributions cut in 2026.

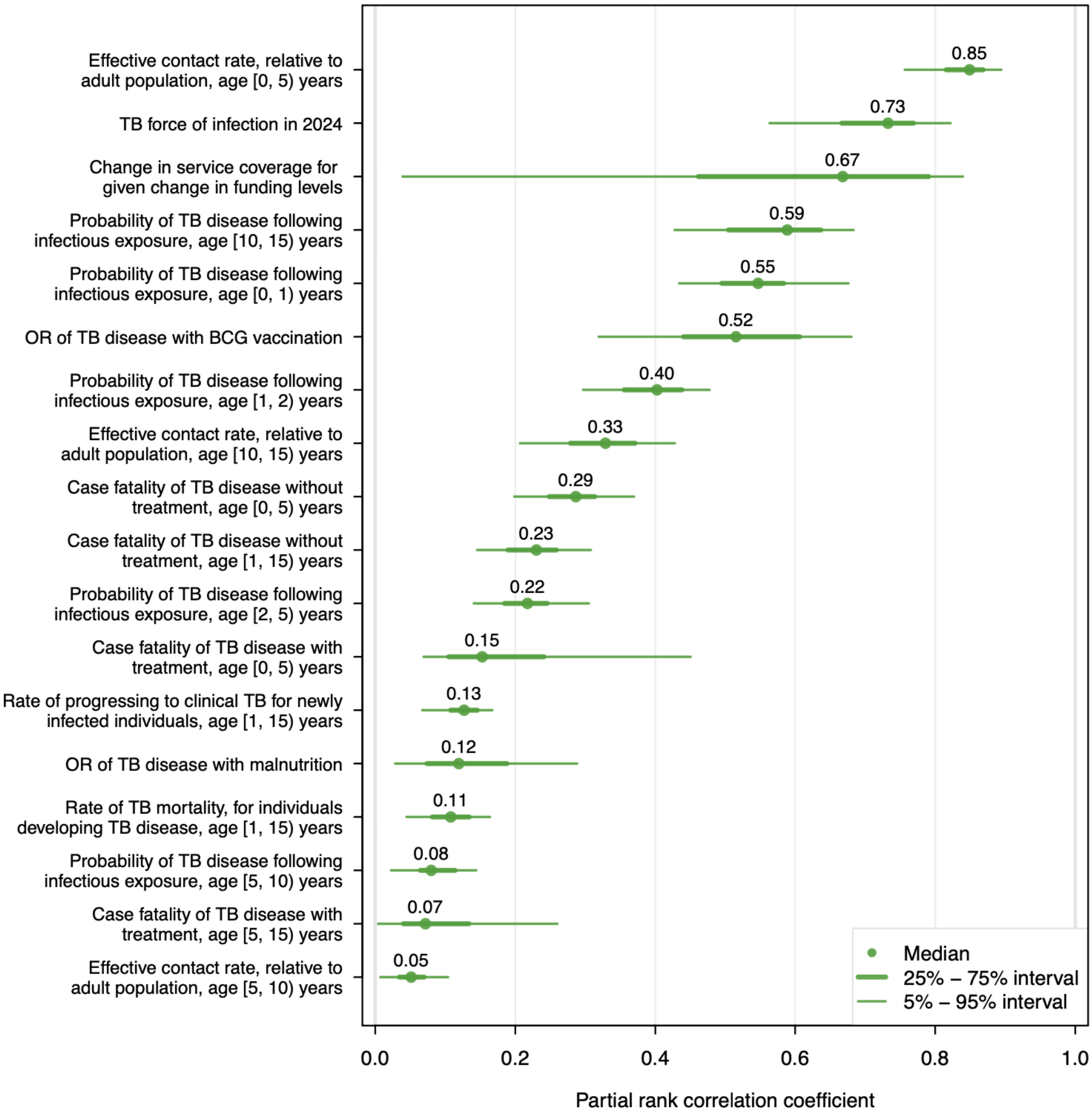

**Figure S8: Partial rank correlation coefficients describing the relationship between individual model parameters and total pediatric TB deaths 2025-2034 under the most extreme funding reduction scenario compared to continued funding.**

OR = odds ratio. BCG = Bacillus Calmette-Guérin. Scenario represents 100% of US bilateral funding cut in 2025, plus 100% of US and 50% of non-US Global Fund contributions cut in 2026 (Scenario 4). Plot shows partial rank correlation coefficients (PRCCs) for parameters with median absolute PRCC>0.05. PRCCs calculated for each country individually, plotted values represent quantiles calculated from distribution of country results for each parameter.

| | Pediatric TB cases | | | Pediatric TB deaths | | |
|---|---|---|---|---|---|---|
| | 2025-2029 | 2030-2034 | Whole period | 2025-2029 | 2030-2034 | Whole period |
| *Absolute increase (millions)* | | | | | | |
| Low impact scenario: US bilateral funding cut during 2025 then restored to full funding | 0.19 (0.14, 0.24) | 0.14 (0.10, 0.18) | 0.33 (0.25, 0.42) | 0.026 (0.019, 0.034) | 0.013 (0.010, 0.017) | 0.039 (0.029, 0.051) |
| High impact scenario: as in Scenario 4, but with HIV and TB services reductions proportional to funding reductions | 2.7 (2.2, 3.3) | 14 (12, 18) | 17 (14, 21) | 0.73 (0.60, 0.90) | 2.9 (2.3, 3.6) | 3.6 (2.9, 4.5) |
| *Relative increase (%)* | | | | | | |
| Low impact scenario: US bilateral funding cut during 2025 then restored to full funding | 3.2 (2.6, 3.9) | 2.6 (2.0, 3.1) | 2.9 (2.3, 3.5) | 4.5 (3.7, 5.6) | 2.4 (1.9, 2.9) | 3.5 (2.8, 4.3) |
| High impact scenario: as in Scenario 4, but with HIV and TB services reductions proportional to funding reductions | 46 (41, 52) | 264 (225, 302) | 151 (129, 172) | 130 (112, 148) | 543 (456, 639) | 330 (279, 385) |

**Table S6: Additional pediatric TB cases and deaths in low- and middle-income countries projected under low impact and high impact scenarios, as compared to continued funding, 2025-2034.**
